# A Scoping Review of ‘Pacing’ for Management of Myalgic Encephalomyelitis/Chronic Fatigue Syndrome (ME/CFS): Lessons Learned for the Long COVID Pandemic

**DOI:** 10.1101/2023.08.10.23293935

**Authors:** Nilihan E.M. Sanal-Hayes, Marie Mclaughlin, Lawrence D. Hayes, Jacqueline L. Mair, Jane Ormerod, David Carless, Natalie Hilliard, Rachel Meach, Joanne Ingram, Nicholas F. Sculthorpe

**Affiliations:** Sport and Physical Activity Research Institute, School of Health and Life Sciences, University of the West of Scotland, Glasgow, UK; Future Health Technologies, Singapore-ETH Centre, Campus for Research Excellence and Technological Enterprise (CREATE), Singapore; Saw Swee Hock School of Public Health, National University of Singapore, Singapore; Long COVID Scotland, 12 Kemnay Place, Aberdeen, UK; Physios for ME, Online, UK; School of Education and Social Sciences, University of the West of Scotland, Glasgow, UK

**Keywords:** Pacing, Myalgic Encephalomyelitis, Chronic Fatigue Syndrome, Post-Exertional Malaise

## Abstract

**Background:** Controversy over treatment for people with myalgic encephalomyelitis/chronic fatigue syndrome (ME/CFS) is a barrier to appropriate treatment. Energy management or *pacing* is a prominent coping strategy for people with ME/CFS that involves regulating activity to avoid post exertional malaise (PEM), the worsening of symptoms after an activity. Until now, characteristics of pacing, and the effects on patients’ symptoms had not been systematically reviewed. This is problematic as the most common approach to pacing, pacing prescription, and the pooled efficacy of pacing was unknown. Collating evidence may help advise those suffering with similar symptoms, including long COVID, as practitioners would be better informed on methodological approaches to adopt, pacing implementation, and expected outcomes.

**Objectives:** In this scoping review of the literature, we aggregated type of, and outcomes of, pacing in people with ME/CFS.

**Eligibility criteria:** Original investigations concerning pacing were considered in participants with ME/CFS.

**Sources of evidence:** Six electronic databases (PubMed, Scholar, ScienceDirect, Scopus, Web of Science and the Cochrane Central Register of Controlled Trials [CENTRAL]) were searched; and websites MEPedia, Action for ME, and ME Action were also searched for grey literature.

**Methods:** A scoping review was conducted. Review selection and characterisation was performed by two independent reviewers using pretested forms.

**Results:** Authors reviewed 177 titles and abstracts, resulting in included 17 studies: three randomised control trials (RCTs); one uncontrolled trial; one interventional case series; one retrospective observational study; two prospective observational studies; four cross-sectional observational studies; and five cross-sectional analytical studies. Studies included variable designs, durations, and outcome measures. In terms of pacing administration, studies used educational sessions and diaries for activity monitoring. Eleven studies reported benefits of pacing, four studies reported no effect, and two studies reported a detrimental effect in comparison to the control group.

**Conclusions:** Highly variable study designs and outcome measures, allied to poor to fair methodological quality resulted in heterogenous findings and highlights the requirement for more research examining pacing. Looking to the long COVID pandemic, future studies should be RCTs utilising objectively quantified digitised pacing, over a longer duration of examination, using the core outcome set for patient reported outcome measures.

## 1 Introduction

### 1.1 Rationale

Post-viral illness occurs when individuals experience an extended period of feeling unwell after a viral infection [1–3]. While post-viral illness is generally a non-specific condition with a constellation of symptoms that may be experienced, fatigue is amongst the most commonly reported [4–6]. For example, our recent systematic review found there was up to 94% prevalence of fatigue in people following acute COVID-19 infection [3]. The increasing prevalence of long COVID has generated renewed interest in symptomology and time-course of post-viral fatigue, with PubMed reporting 72 articles related to “post-viral fatigue” between 2020 and 2022, but less than five for every year since 1990.

As the coronavirus pandemic developed, it became clear that a significant proportion of the population experienced symptoms which persisted beyond the initial viral infection, meeting the definition of a post-viral illness. Current estimates suggest one in eight people develop long COVID [7]and its symptomatology has repeatedly been suggested to overlap with clinical demonstrations of myalgic encephalomyelitis/chronic fatigue syndrome (ME/CFS). In a study by Wong and Weitzer [8], long COVID symptoms from 21 studies were compared to a list of ME/CFS symptoms. Of the 29 known ME/CFS symptoms the authors reported that 25 (86%) were reported in at least one long COVID study suggesting significant similarities. Sukocheva et al. [9] reported that long COVID included changes in immune, cardiovascular, metabolic, gastrointestinal, nervous and autonomic systems. When observed from a pathological stance, this list of symptoms are shared with, or are similar to, the symptoms patients with ME/CFS describe [10]. In fact, a recent article reported 43% of people with long COVID are diagnosed with ME/CFS [10], evidencing the analogous symptom loads.

A striking commonality between long COVID and similar conditions such as ME/CFS is the worsening of symptoms including fatigue, pain, cognitive difficulties, sore throat, and/or swollen lymph nodes following exertion. Termed *post exertional malaise* (PEM) [11–14], lasting from hours to several days, it is arguably one of the most debilitating side effects experienced by those with ME/CFS [13–15]. PEM is associated with considerably reduced quality of life amongst those with ME/CFS, with reduced ability to perform activities of daily living, leading to restraints on social and family life, mental health comorbidities such as depression and anxiety, and devastating employment and financial consequences [16–19]. At present, there is no cure or pharmacological treatments for PEM, and therefore, effective symptom management strategies are required. This may be in part because the triggers of PEM are poorly understood, and there is little evidence for what causes PEM, beyond anecdotal evidence. The most common approach to manage PEM is to incorporate activity pacing into the day-to-day lives of those with ME/CFS with the intention of reducing the frequency of severity of bouts of PEM [20]. Pacing is defined as an approach where patients are encouraged to be as active as possible within the limits imposed by the illness [20–22]. In practice, pacing requires individuals to determine a level at which they can function but which does not lead to a marked increase in fatigue and other symptoms [23,24].

Although long COVID is a new condition [3,11], the available evidence suggests substantial overlap with the symptoms of conditions such as ME/CFS and it is therefore pragmatic to consider the utility of management strategies (such as pacing) used in ME/CFS for people with long COVID. In fact, a recent Delphi study recommended that management of long COVID should incorporate careful pacing to avoid PEM relapse [25]. This position was enforced by a multidisciplinary consensus statement considering treatment of fatigue in long COVID, recommending energy conservation strategies (including pacing) for people with long COVID [26]. Given the estimated >2 million individuals who have experienced long COVID in the UK alone [27–29], there is an urgent need for evidence-based public health strategies. In this context, it seems pragmatic to borrow from the ME/CFS literature.

From a historical perspective, the 2007 NICE guidelines for people with ME/CFS advised both cognitive behavioural therapy (CBT) and graded exercise therapy (GET) should be offered to people with ME/CFS [30]. As of the 2021 update, NICE guidelines for people with ME/CFS do not advise CBT or GET, and the only recommended management strategy is pacing [31]. In the years between changes to these guidelines, the landmark PACE trial [32] was published in 2011. This large, randomised control trial (RCT; n=639) compared pacing with CBT and reported GET and CBT were more effective than pacing for improving symptoms. Yet, this study has come under considerable criticism from patient groups and clinicians alike [33–36]. This may partly explain why NICE do not advise CBT or GET as of 2021, and only recommend pacing for symptom management people with ME/CFS [31]. There has been some controversy over best treatment for people with ME/CFS in the literature and support groups, potentially amplified by the ambiguity of evidence for pacing efficacy and how pacing should be implemented. As such, before pacing can be advised for people with long COVID, it is imperative previous literature concerning pacing is systematically reviewed. This is because a consensus is needed within the literature for implementing pacing so practitioners treating people with ME/CFS or long COVID can do so effectively. A lack of agreement in pacing implementation is a barrier to adoption for both practitioners and patients. Despite a number of systematic reviews concerning pharmacological interventions or cognitive behavioural therapy in people with ME/CFS [33,37,38], to date, there are no systematic reviews concerning pacing.

Despite the widespread use of pacing, the literature base is limited and includes clinical commentaries, case studies, case series, and few randomised control trials. Consequently, while a comprehensive review of the effects of pacing in ME/CFS is an essential tool to guide symptom management advice, the available literature means that effective pooling of data is not feasible [39] and therefore, a traditional systematic review and meta-analysis, with a tightly focussed research question would be premature [40]. Consequently, we elected to undertake a scoping review. This approach retains the systematic approach to literature searching but aims to map out the current state of the research [40]. Using the framework of Arksey and O’Malley [41], a scoping review aims to use a broad set of search terms and include a wide range of study designs and methods (in contrast to a systematic review [41]). This approach, has the benefit of clarifying key concepts, surveying current data collection approaches, and identifying critical knowledge gaps.

### 1.2 Objectives

We aimed to provide an overview of existing literature concerning pacing in ME/CFS. Our three specific objectives of this scoping review were to (1) conduct a systematic search of the published literature concerning ME/CFS and pacing, (2) map characteristics and methodologies used, and (3) provide recommendations for the advancement of the research area.

## 2 Methods

### 2.1 Protocol and Registration

The review was conducted and reported according to the Preferred Reporting Items for Systematic Reviews and Meta-Analyses extension for scoping reviews (PRISMA-ScR) guidelines [42] and the five-stage framework outlined in Arksey and O’Malley [41]. Registration is not recommended for scoping reviews.

### 2.2 Eligibility criteria

Studies that met the following criteria were included in this review: (1) published as a full-text manuscript; (2) not a review; (3) participants with ME/CFS; (4) studies employed a pacing intervention or retrospective analysis of pacing or a case study of pacing. Studies utilising sub-analysis of the pacing, graded activity, and cognitive behaviour therapy: a randomised evaluation (PACE) trial were included as these have different outcome measures and, as this is not a meta-analysis, this will not influence effect size estimates. Additionally, due to the paucity of evidence, grey literature has also been included in this review.

### 2.3 Search Strategy

The search strategy consisted of a combination of free-text and MeSH terms relating to ME/CFS and pacing, which were developed through an examination of published original literature and review articles. Example search terms for PubMed included: ‘ME/CFS’ OR ‘ME’ OR ‘CFS’ OR ‘chronic fatigue syndrome’ OR ‘PEM’ OR ‘post exertional malaise’ OR ‘pene’ OR ‘post-exertion neurogenic exhaust’ AND ‘pacing’ OR ‘adaptive pacing’. The search was performed within title/abstract. Full search terms can be found in **Appendix 1**.

### 2.4 Information sources

Six electronic databases [PubMed, Scholar, ScienceDirect, Scopus, Web of Science, and the Cochrane Central Register of Controlled Trials (CENTRAL)] were searched to identify original research articles published from the earliest available date up until 02/02/2022. Additional records were identified through reference lists of included studies. ‘Grey literature’ repositories including MEPedia, Action for ME, and ME Action were also searched with the same terms.

### 2.5 Study selection and Data Items

Once each database search was completed and manuscripts were sourced, all studies were downloaded into a single reference list (Zotero, version 6.0.23) and duplicates were removed. Titles and abstracts were screened for eligibility by two reviewers independently and discrepancies were resolved through discussion between reviewers. Subsequently, full text papers of potentially relevant studies were retrieved and assessed for eligibility by the same two reviewers independently. Any uncertainty by reviewers was discussed in consensus meetings and resolved by agreement. Data extracted from each study included sample size, participant characteristics, study design, trial registration details, study location, pacing description (type), intervention duration, intervention adherence, outcome variables, and main outcome data. Descriptions were extracted with as much detail as was provided by the authors. Study quality was assessed using the Physiotherapy Evidence Database (PEDro) scale [43,44].

### 2.6 Role of the funding source

The study sponsors had no role in study design, data collection, analysis, or interpretation, nor writing the report, nor submitting the paper for publication.

## 3 Results

### 3.1 Study selection

After the initial database search, 281 records were identified (see Fig. 1). Once duplicates were removed, 177 titles and abstracts were screened for inclusion resulting in 22 studies being retrieved as full text and assessed for eligibility. Of those, five were excluded, and 17 articles remained and were used in the final qualitative synthesis.

**Fig. 1.**
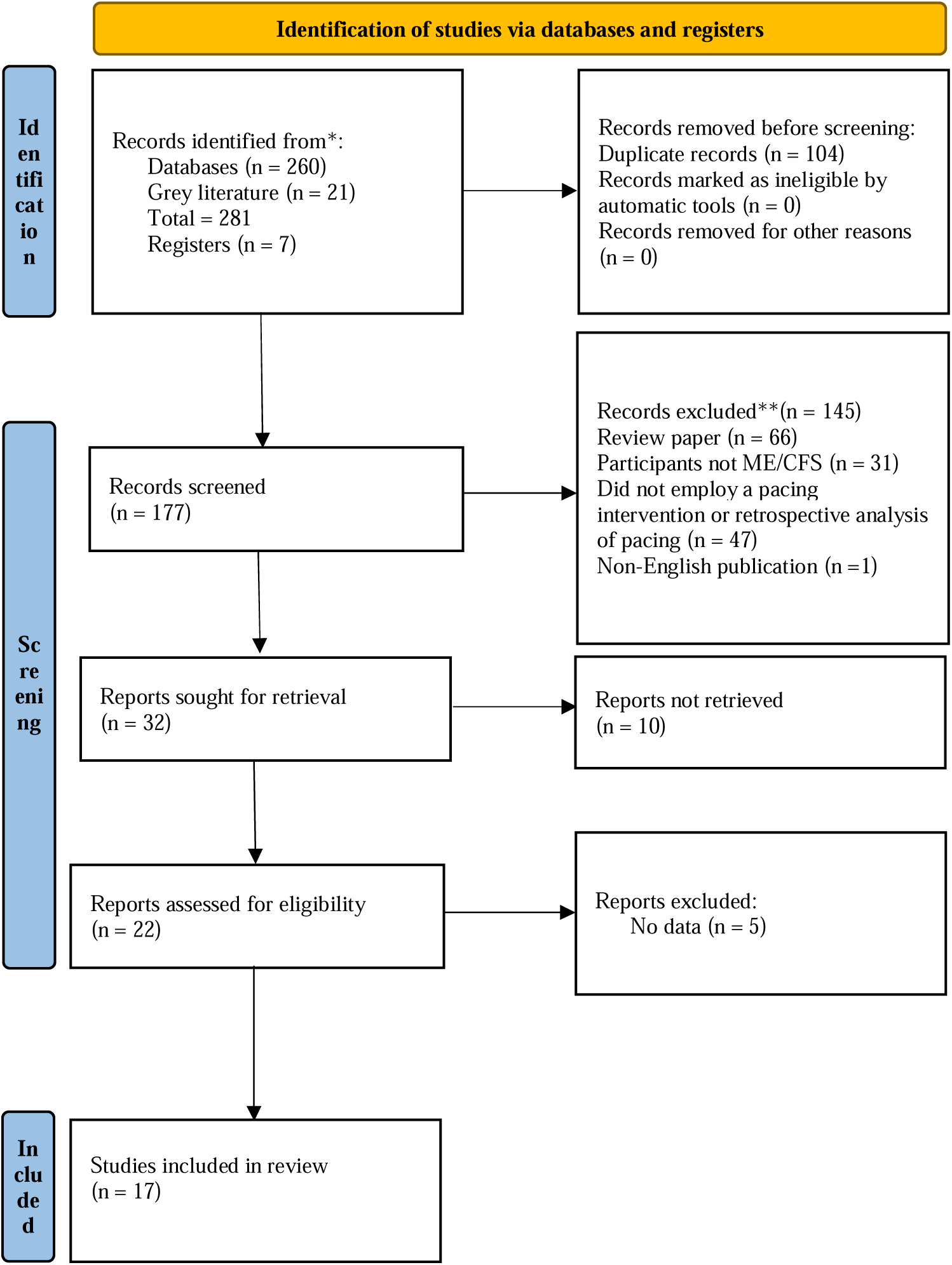
Schematic flow diagram describing exclusions of potential studies and final number of studies. RCT = randomized control trial. CT = controlled trial. UCT = uncontrolled trial.

### 3.2 Study characteristics

Study characteristics are summarised in table 1. Of the 17 studies included, three were randomised control trials (RCTs [32,45,46]); one was an uncontrolled trial [47]; one was a case series [48]; one was a retrospective observational study [49], two were prospective observational studies [50,51]; four were cross-sectional observational studies [22,52,53]; and five were cross-sectional analytical studies [54–58] including sub-analysis of the PACE trial [32,53,56,58]. Seven of the studies were registered trials [32,45–47,53–55]. Diagnostic criteria for ME/CFS are summarised in table 2.

**Table 1.**
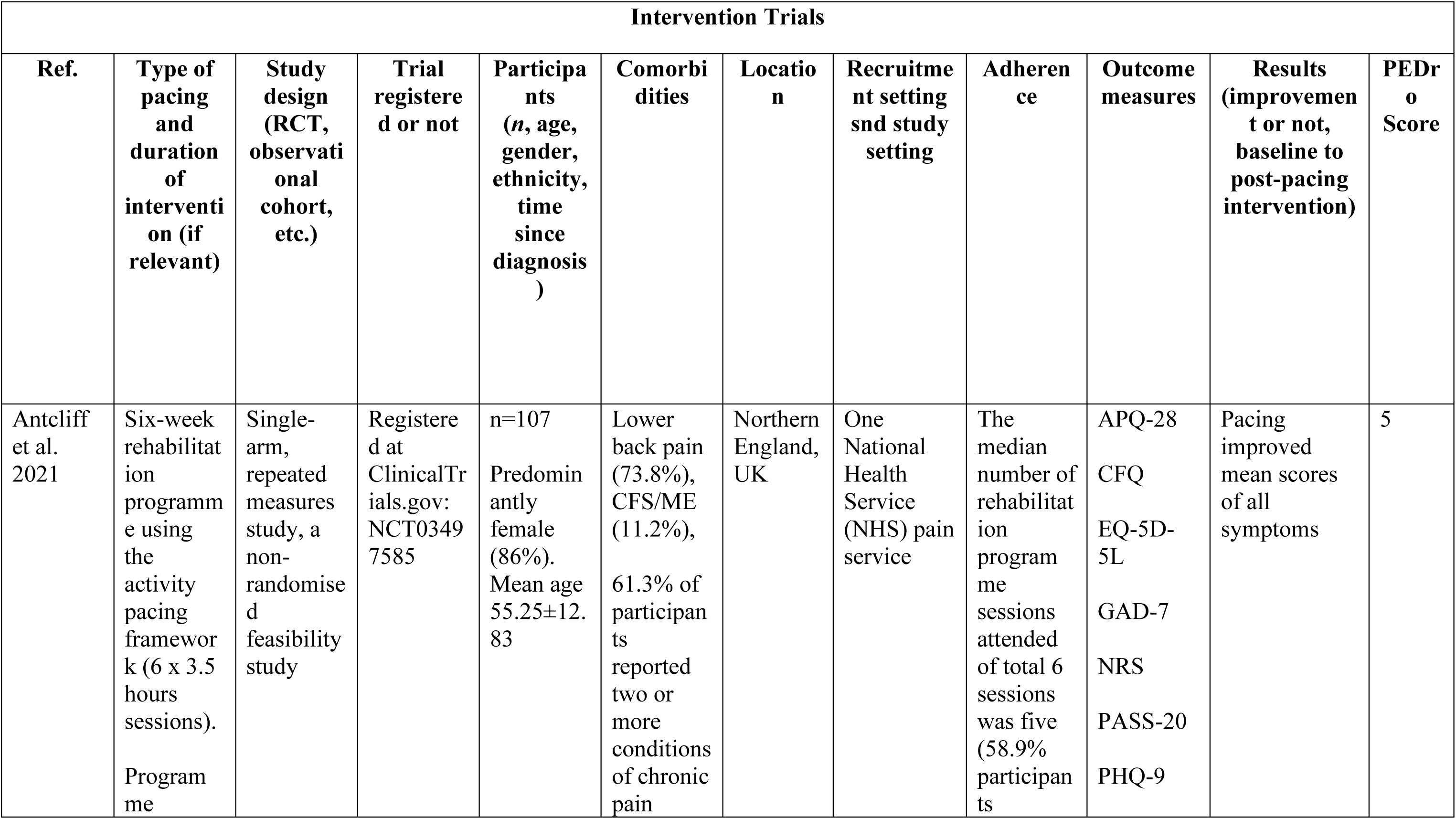
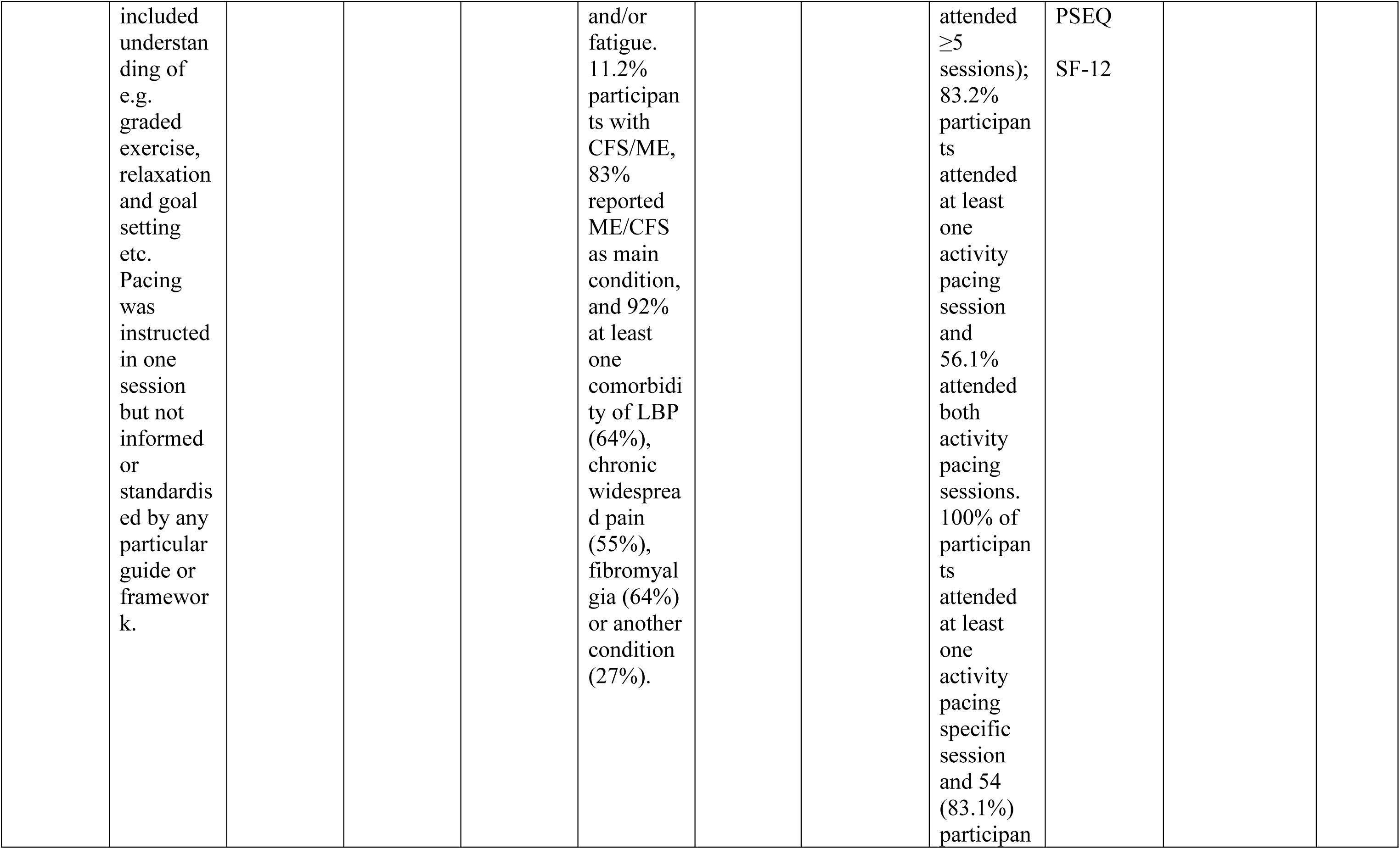
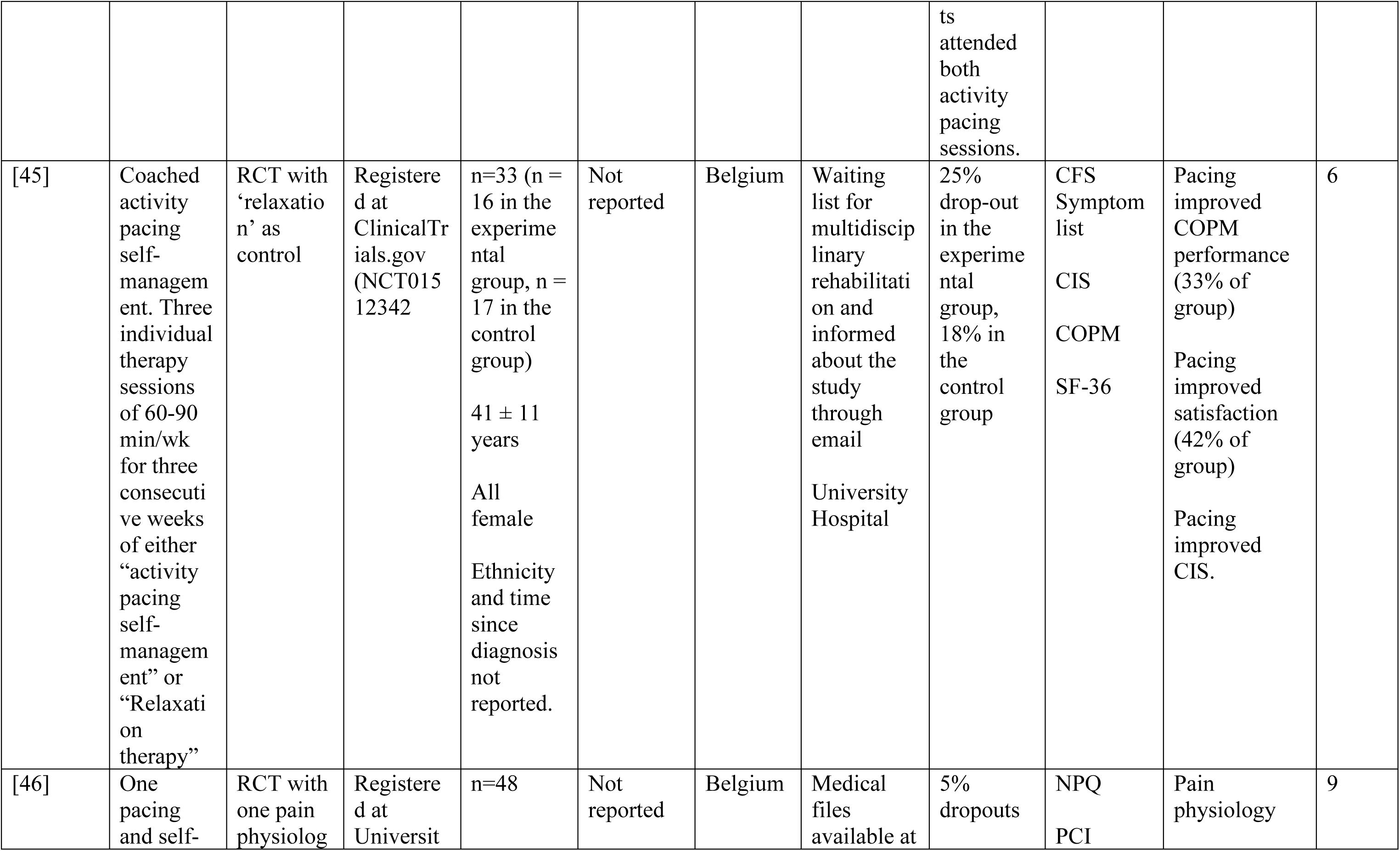
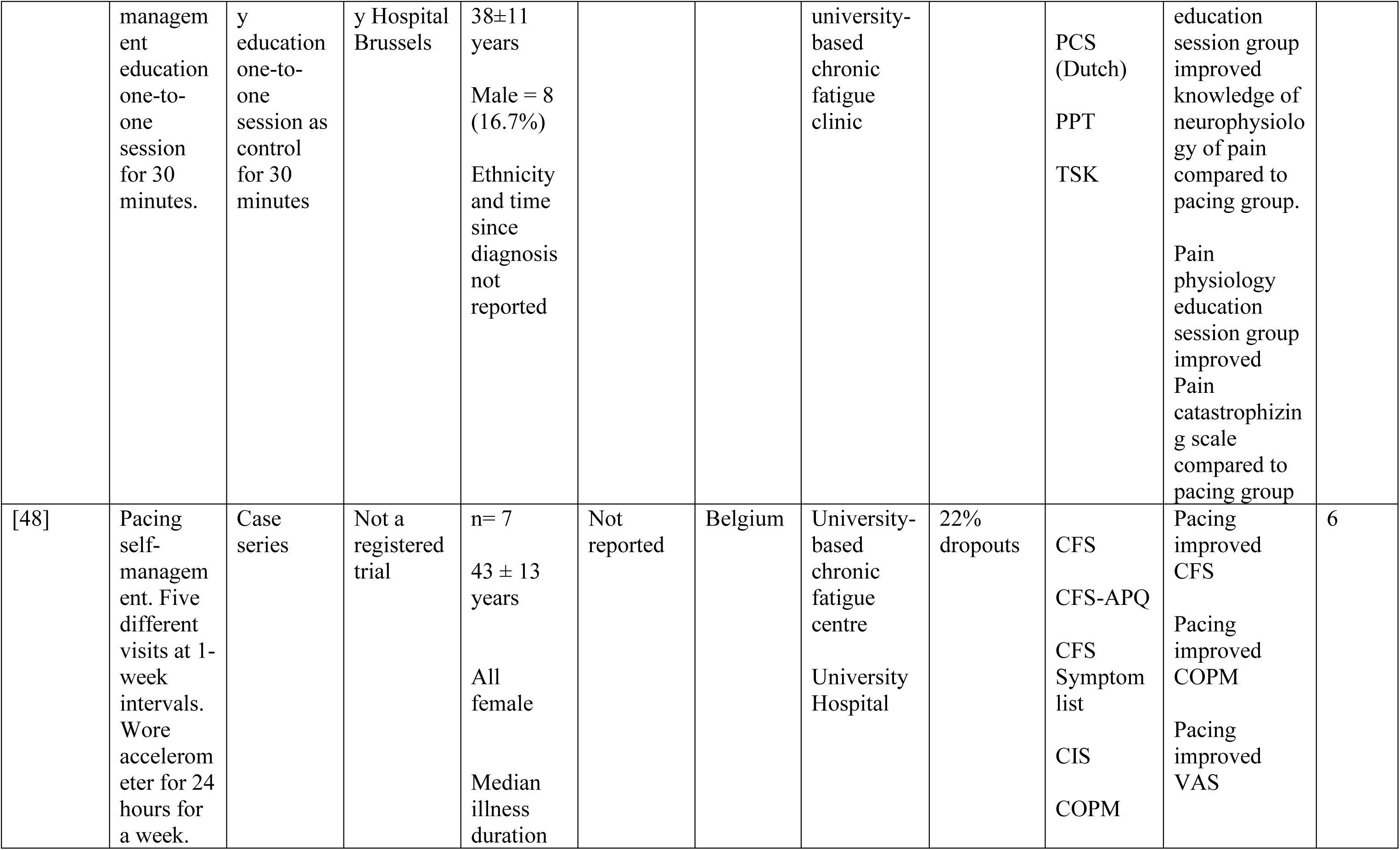
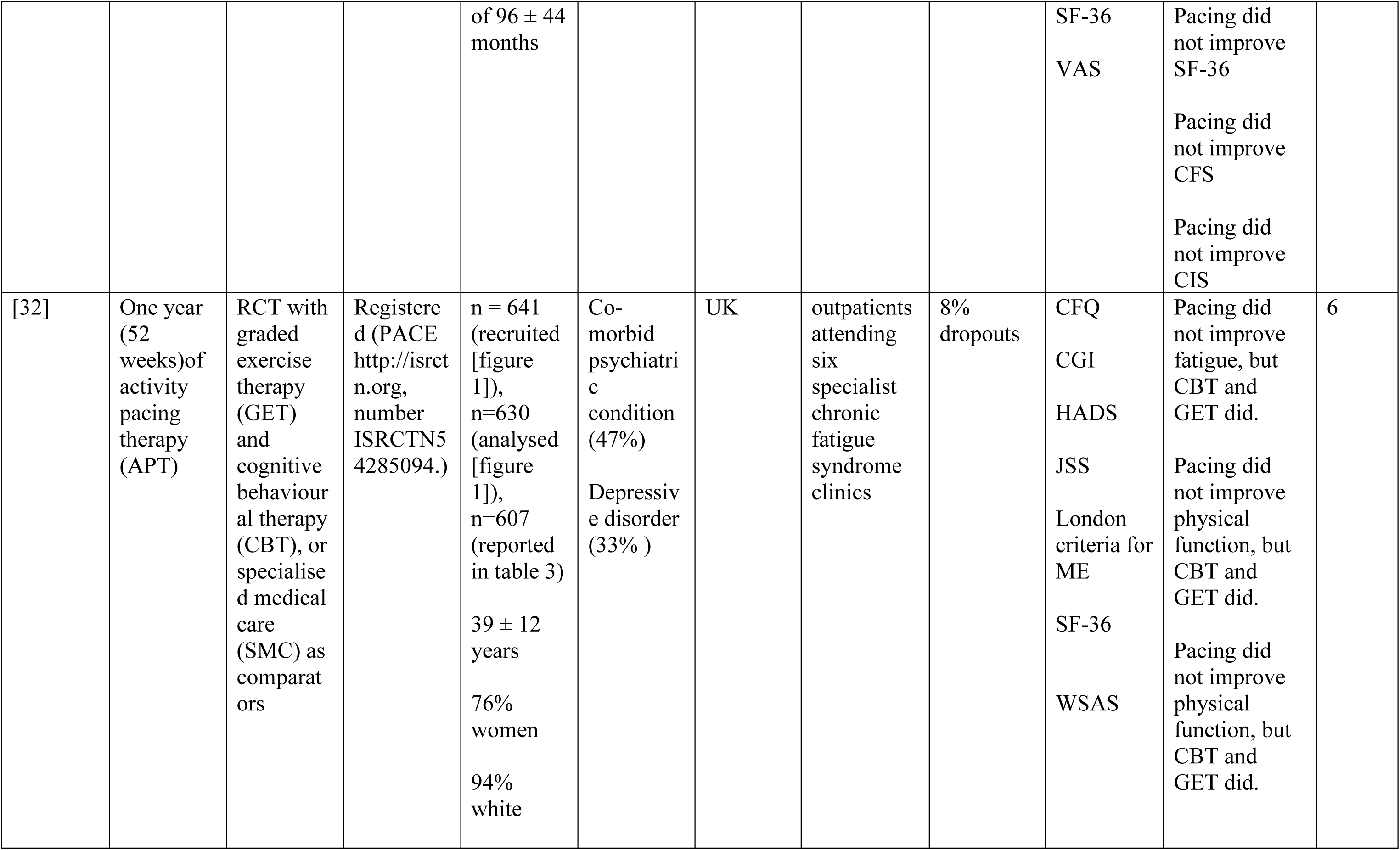
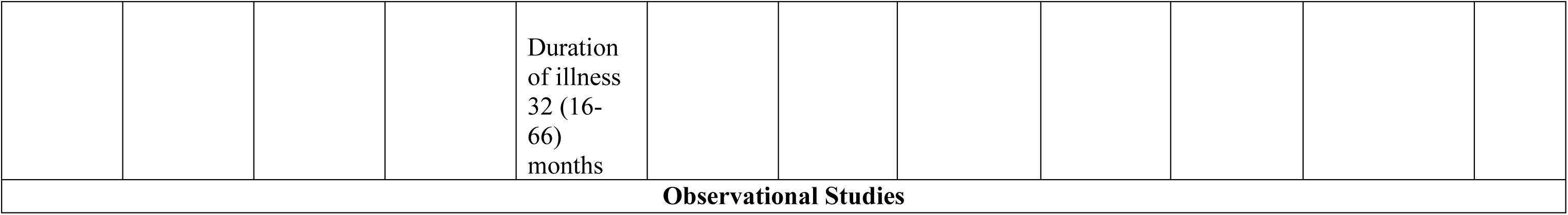
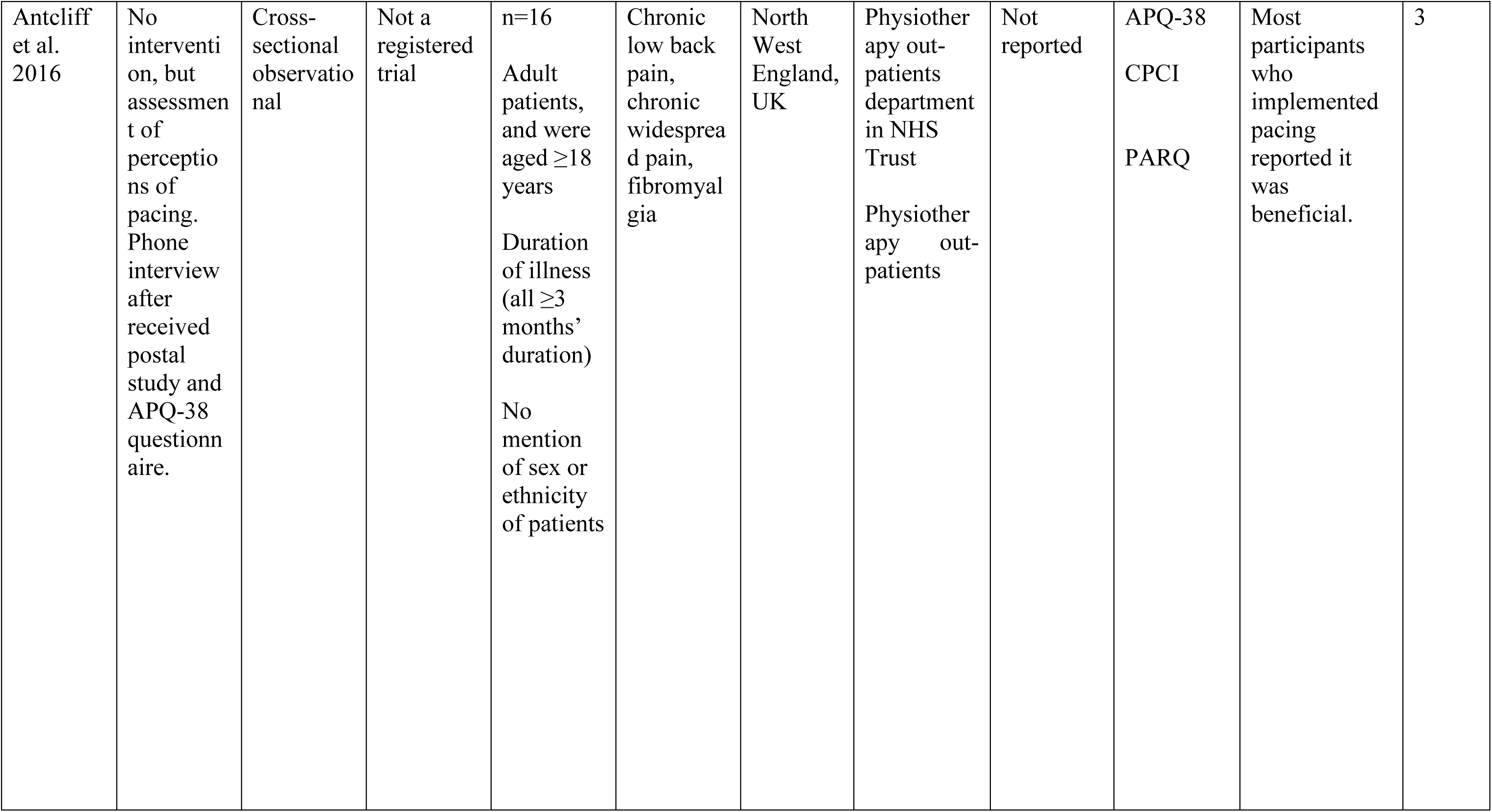
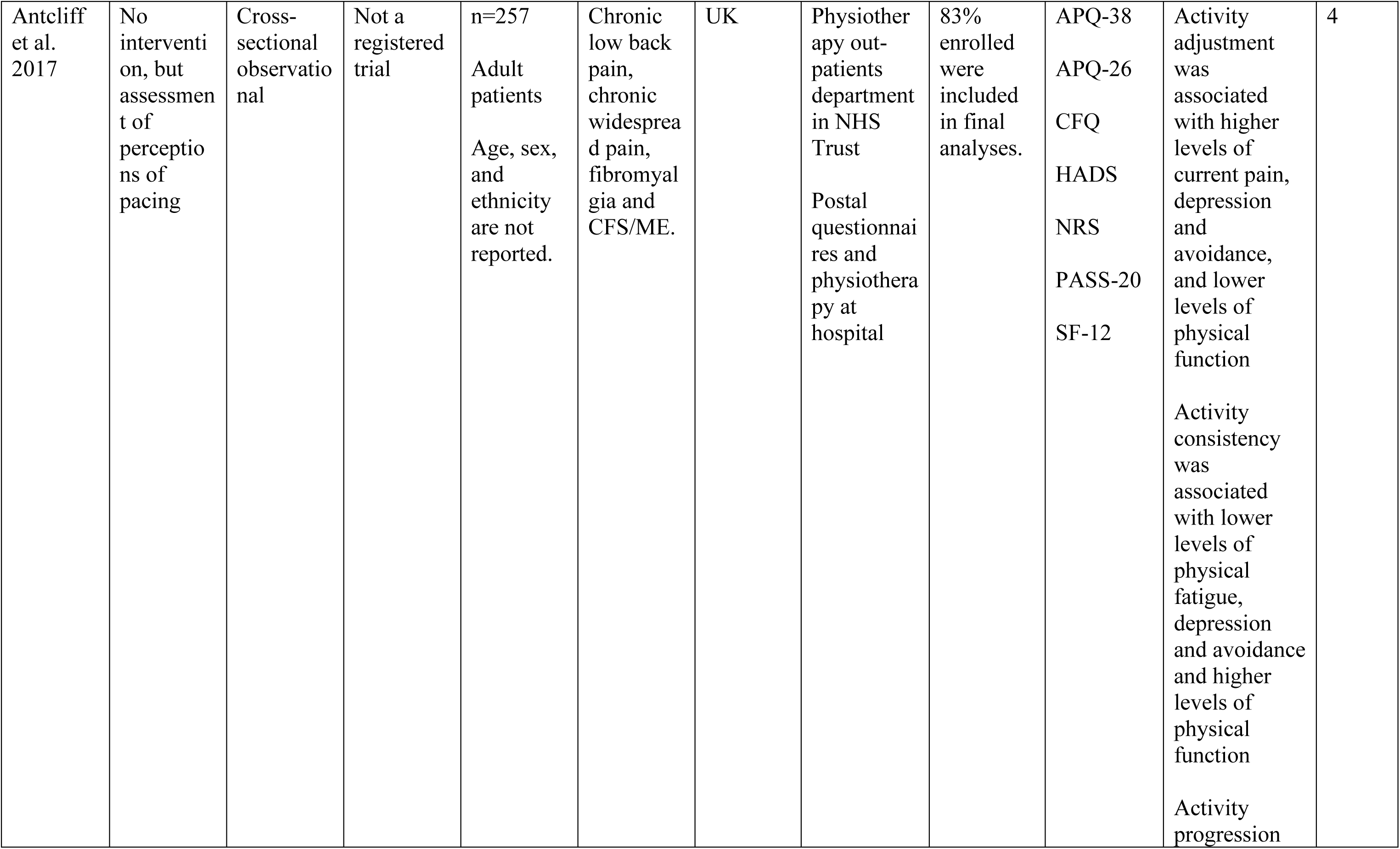
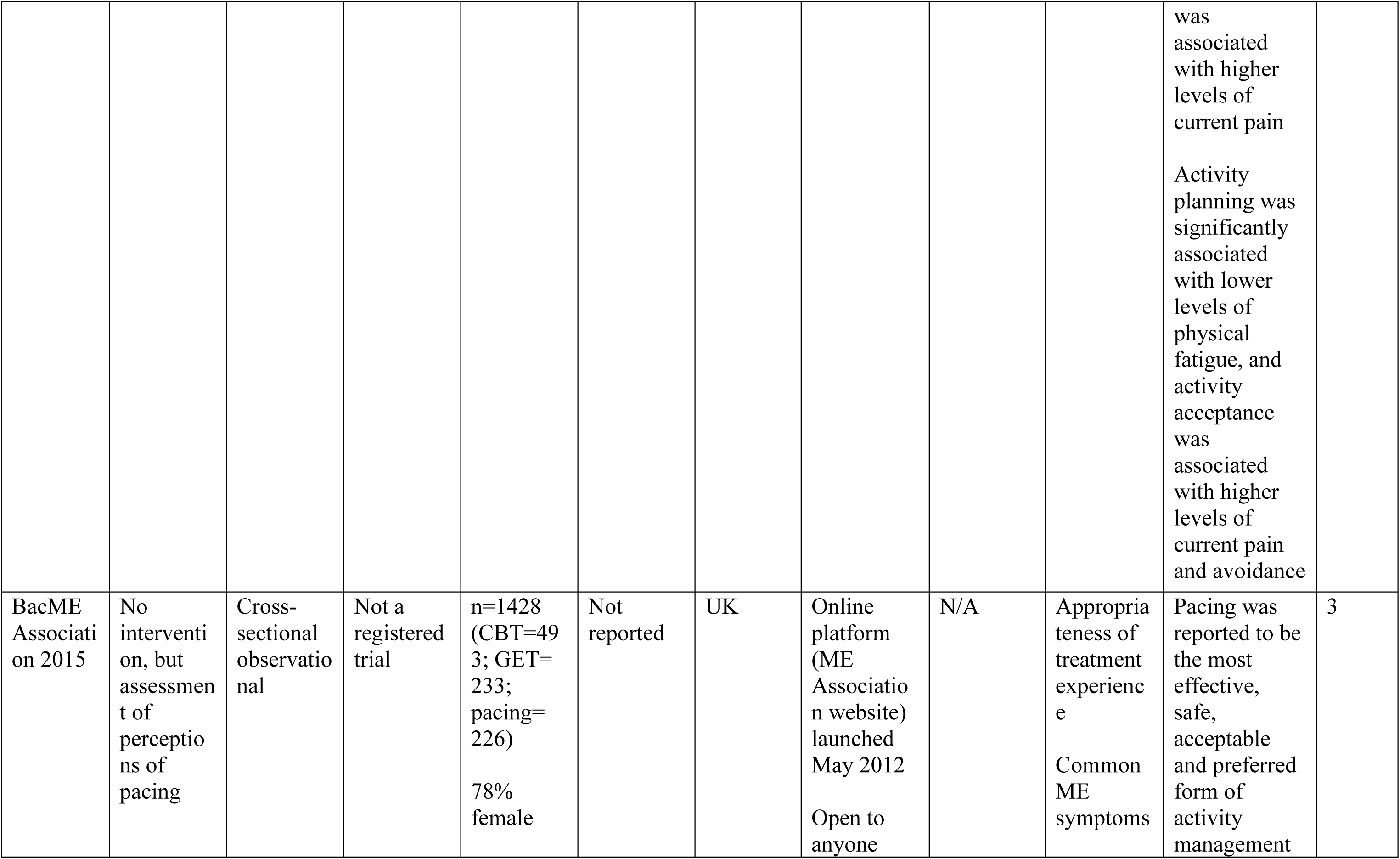
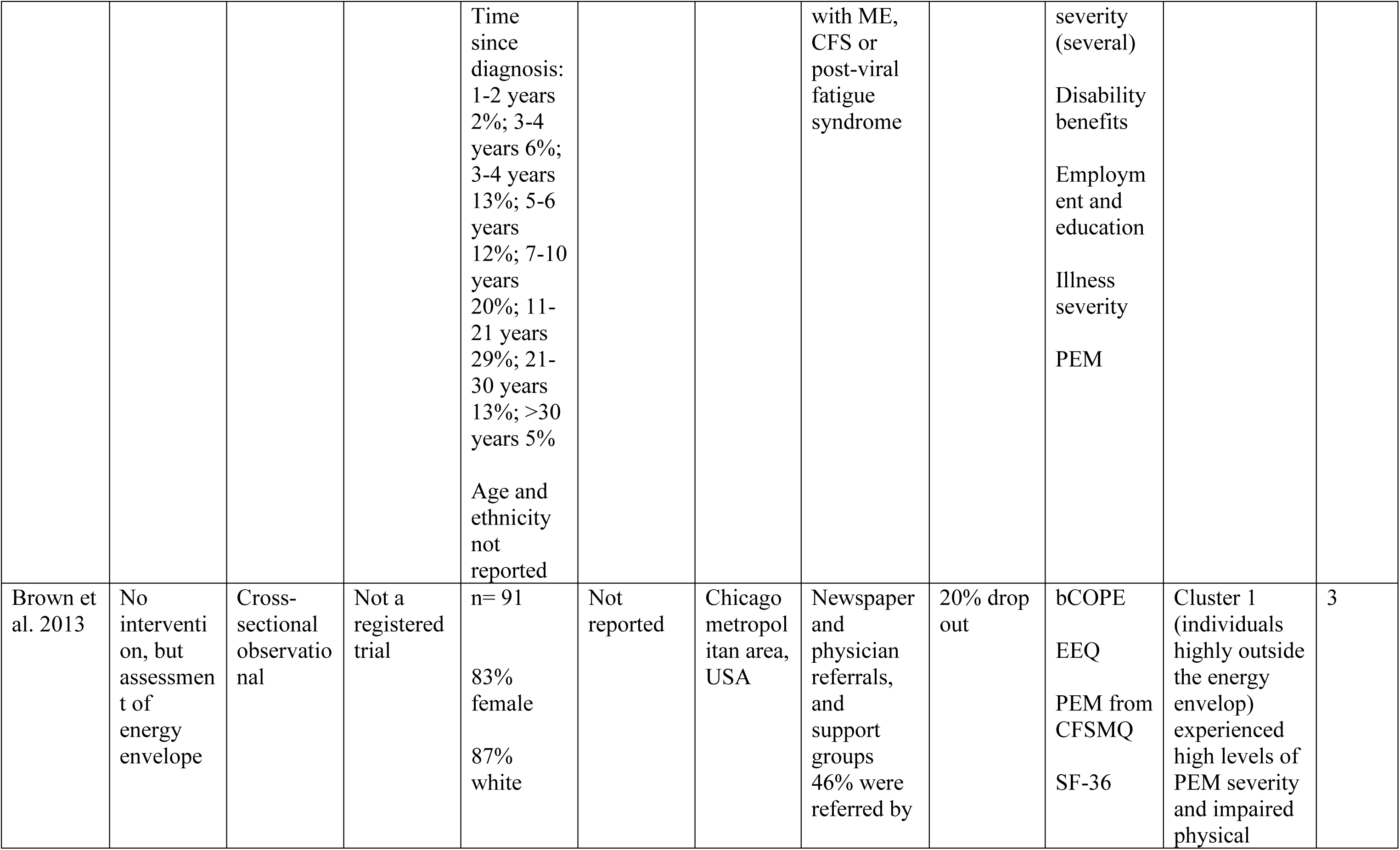
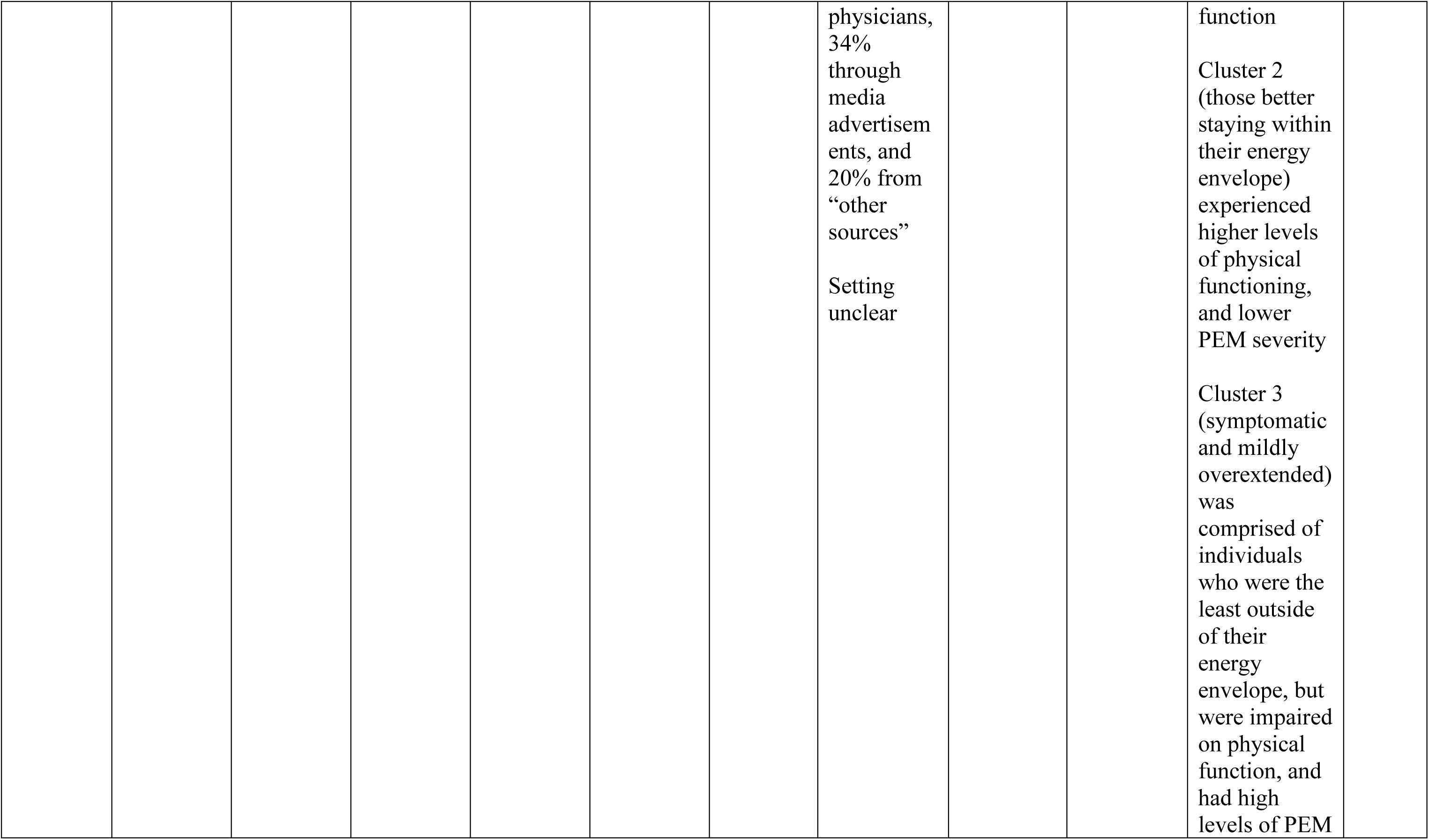
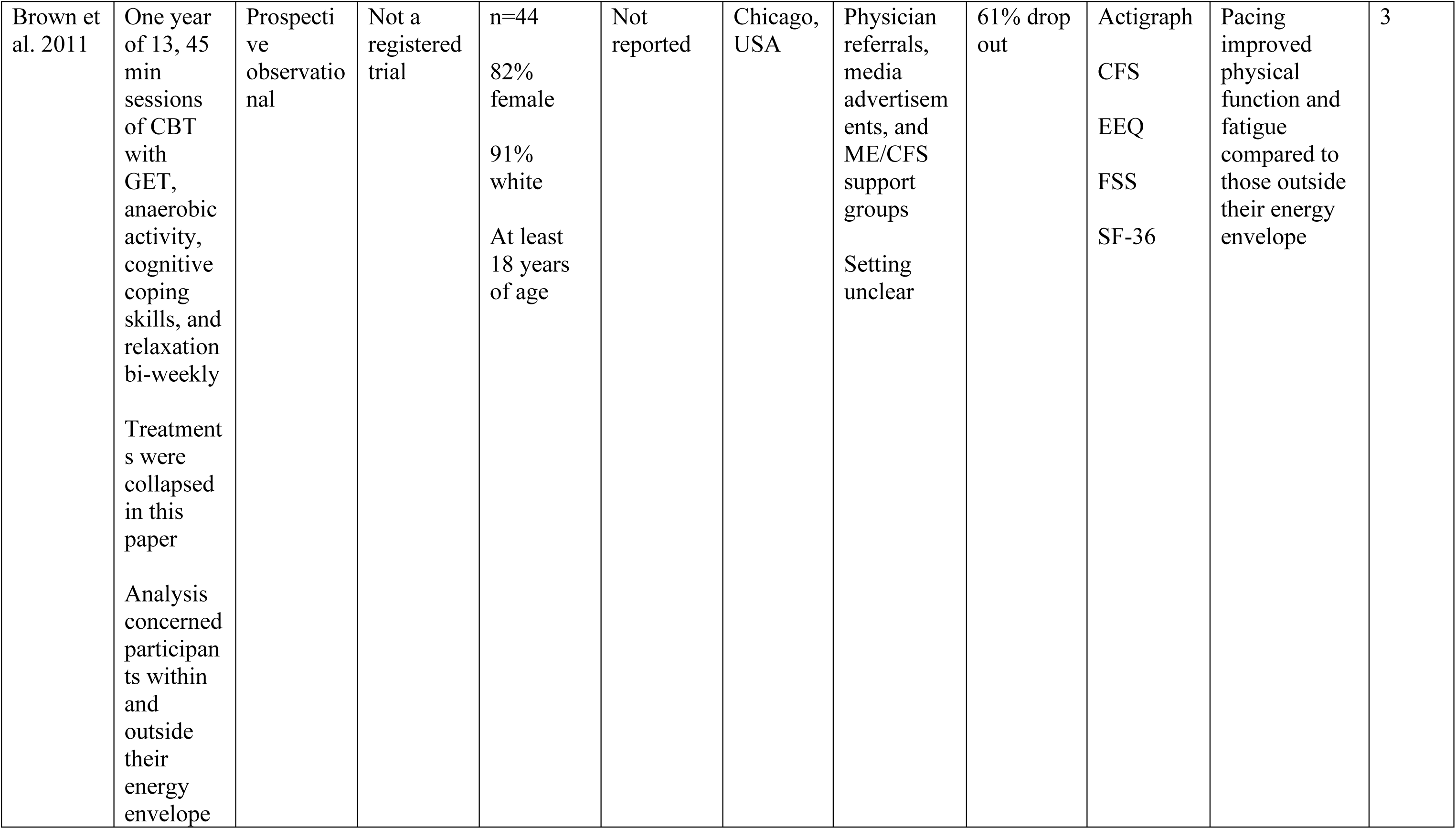
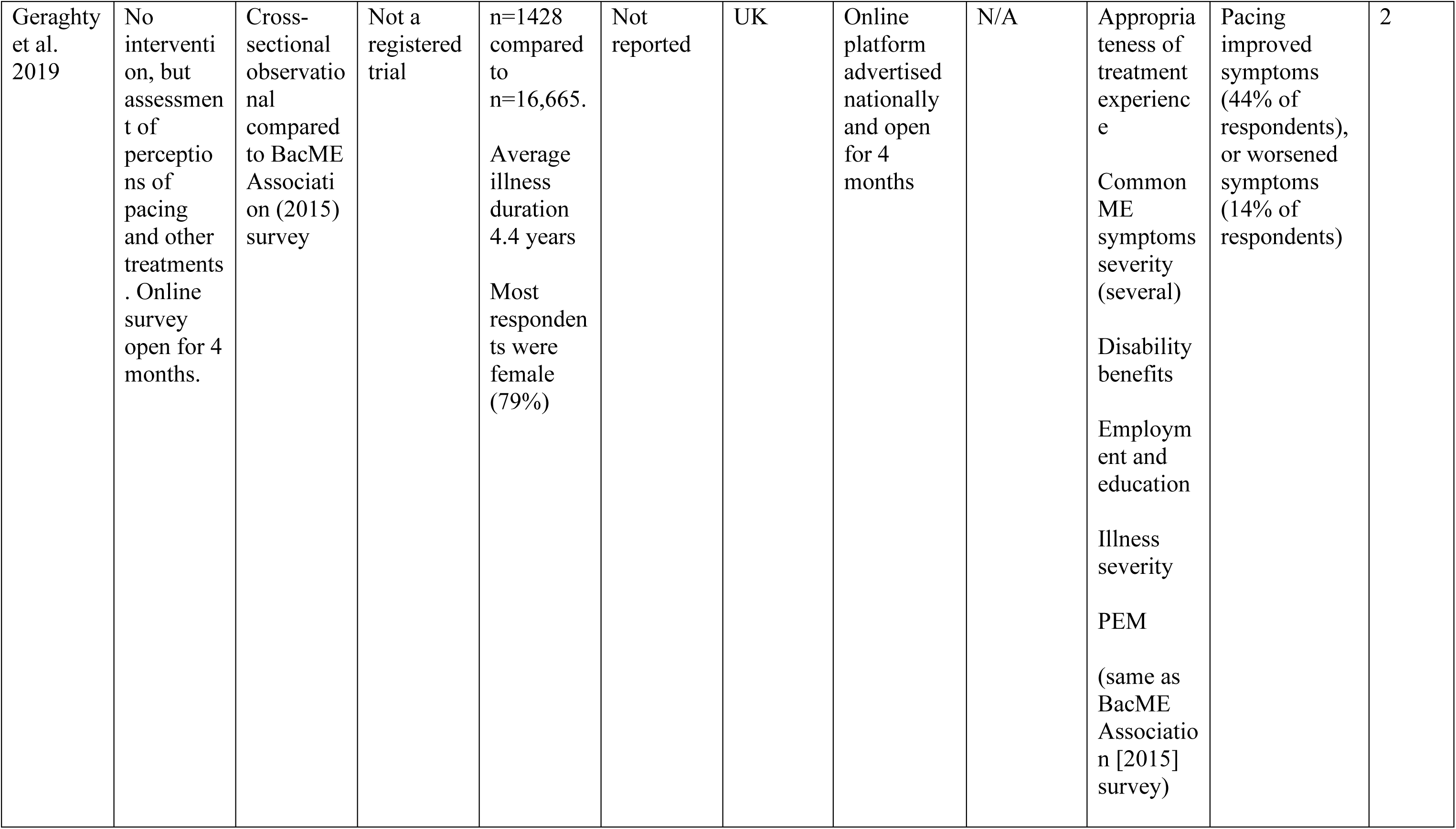
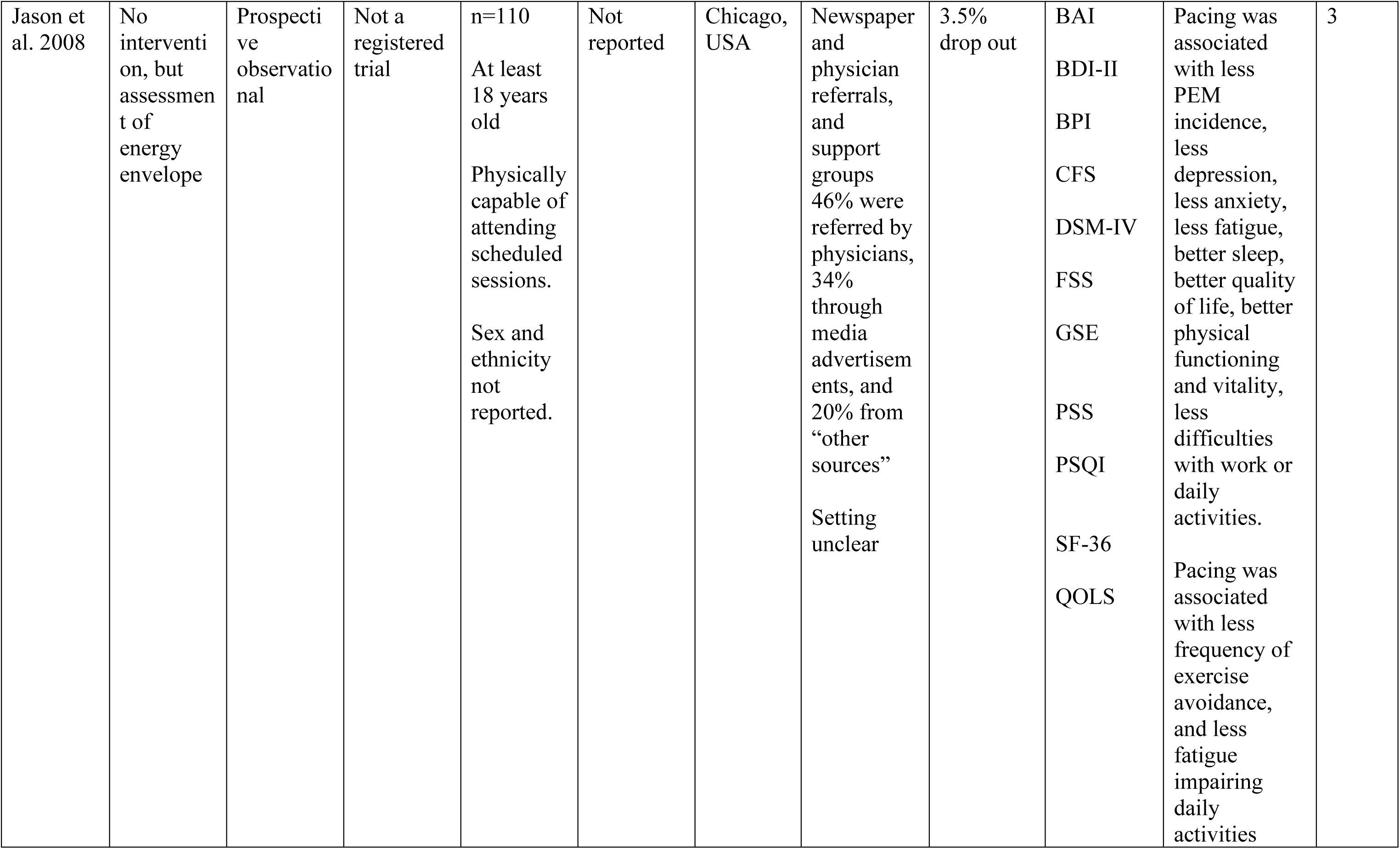
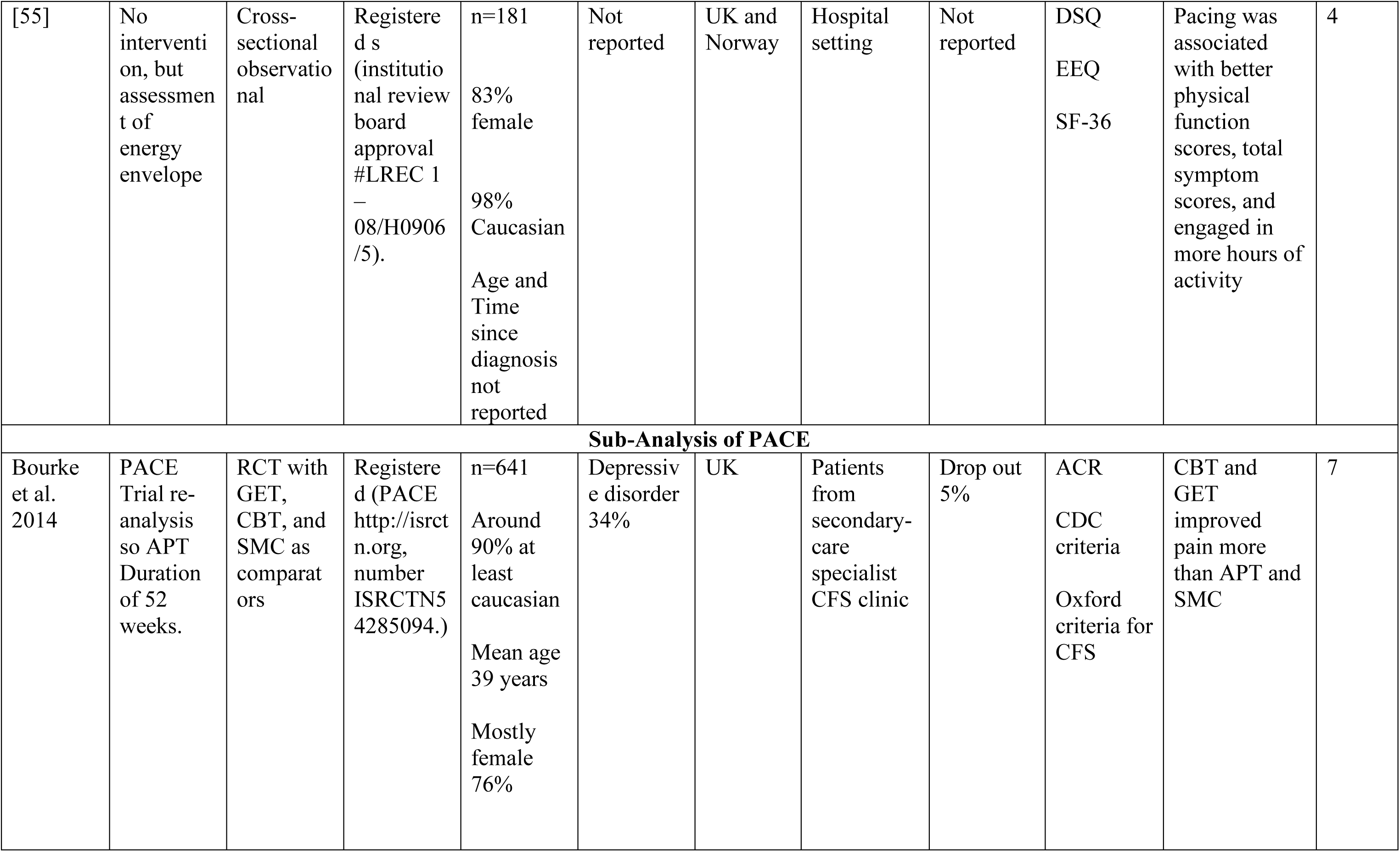
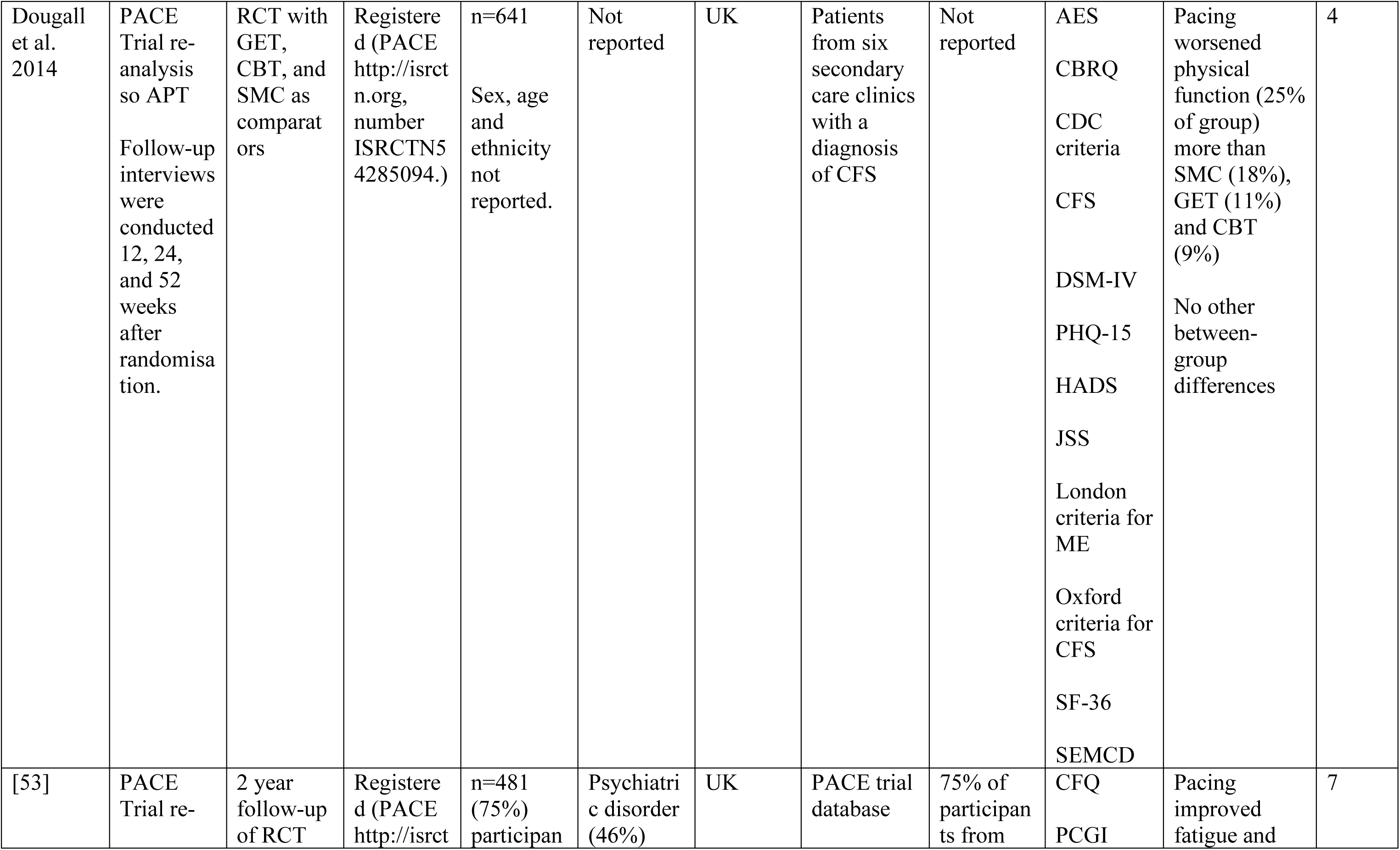
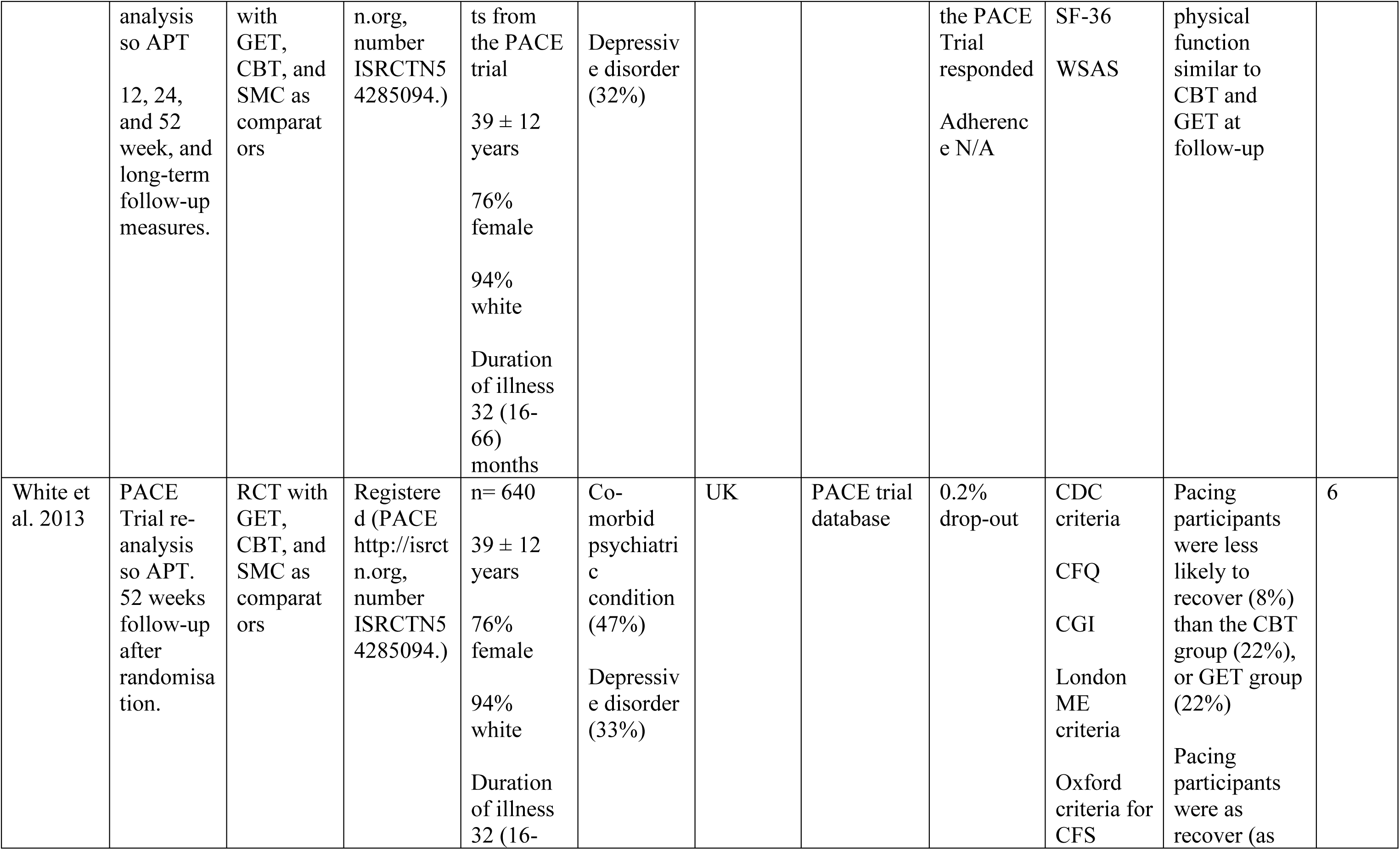
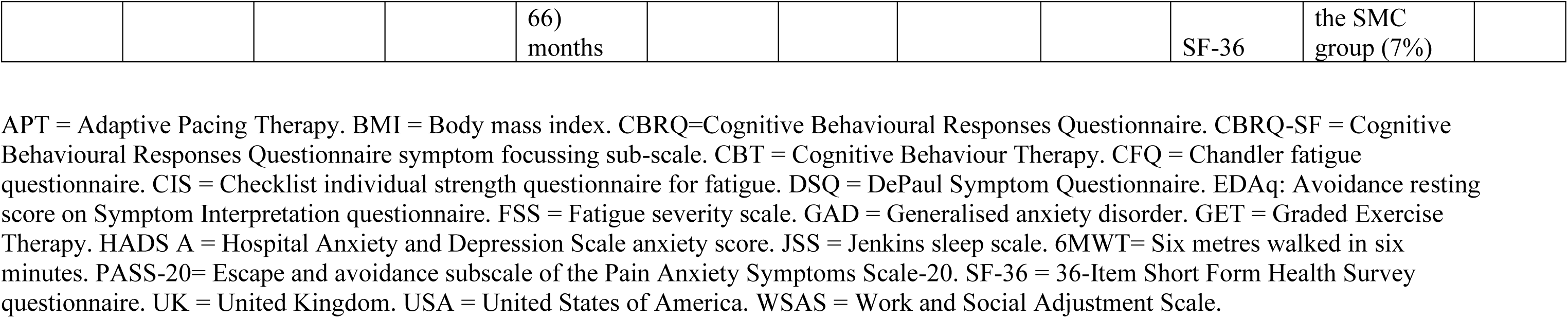
Description of included studies and data sets. RCT = randomised control trial; UCT = uncontrolled trial.

**Table 2.** Diagnostic criteria used in included studies to define ME/CFS (if reported).

| **Study** | **Diagnostic criteria used** |
| --- | --- |
| Antcliff et al., 2016 | No diagnostic criteria included.<br>Participants had diagnosis of chronic low back pain, chronic widespread pain, fibromyalgia and ME/CFS for $\geq 3$ months’. |
| Antcliff et al., 2017 | No diagnostic criteria included.<br>Participants had diagnosis of chronic low back pain, chronic widespread pain, fibromyalgia and ME/CFS for $\geq 3$ months’. |
| Antcliff et al., 2021 | No diagnostic criteria included.<br>Participants had diagnosis of chronic low back pain, chronic widespread pain, fibromyalgia and ME/CFS for $\geq 3$ months’. |
| ME association.,2015 | No diagnostic criteria included.<br>Survey open to anyone who had had ME, CFS, or post-viral fatigue syndrome. |
| Bourke et al., 2014 | **Oxford criteria** (PACE trial sub-analysis). |
| Brown et al., 2011 | **Fukuda criteria.** |
| Brown et al., 2013 | No diagnostic criteria included. |
| Dougall et al., 2014 | **Oxford criteria** (PACE trial sub-analysis). |
| Geraghty et al., 2017 | No diagnostic criteria included.<br>Survey open to anyone who had ME/CFS. According to self-report, 72% received a diagnosis of by a specialist, 22% from a general practitioner (family doctor), and 4.5% from other professional. |
| Jason et al., 2008 | **Fukuda criteria.** |
| Kos et al., 2015 | **Fukuda criteria.** |
| Meeus et al., 2010 | **Fukuda criteria.** |
| Nijs et al., 2009 | **Fukuda criteria.** |
| O’Connor et al., 2019 | **Fukuda criteria.** |
| Sharpe et al., 2015 | **Oxford criteria** (PACE trial sub-analysis). |
| White et al., 2011 | **Oxford criteria** (PACE trial). |
| White et al., 2013 | **Oxford criteria** (PACE trial sub-analysis). |

### 3.3 Types of Pacing

#### 3.3.1 Pacing Interventions

Of the 17 studies included, five implemented their own pacing interventions and will be discussed in this section. Sample sizes ranged from n=7 in an interventional case series [48] to n=641 participants in the largest RCT [32]. The first of these five studies considered an education session on pacing and self-management as the ‘pacing’ group, and a ‘pain physiology education’ group as the control group [46]. Two studies included educational sessions provided by a therapist plus activity monitoring via ActiGraph accelerometers [48] and diaries [45] at baseline and follow-up. In the first of these two studies, Nijs and colleagues [48] implemented a ‘self-management program’ which asked patients to estimate their current physical capabilities prior to commencing an activity and then complete 25-50% less than their perceived energy envelope. They[48] did not include a control group and had a sample size of only n=7. Six years later, the same research group [45] conducted another pacing study which utilised relaxation as a comparator group (n=12 and n=14 in the pacing and relaxation groups, respectively). The pacing group underwent a pacing phase whereby participants again aimed to complete 25-50% less than their perceived energy envelope, followed by a gradual increase in exercise after the pacing phase (the total intervention spanned three weeks, and it is unclear how much was allocated to pacing, and how much to activity increase). Therefore, it could be argued that Kos et al. [45] really assessed pacing followed by a gradual exercise increase as outcome measures were assessed following the graded activity phase. Another pacing intervention delivered weekly educational sessions for six weeks and utilised a standardised rehabilitation programme using the ‘activity pacing framework’ [47] in a single-arm, no comparator group feasibility study. Finally, the PACE trial adopted an adaptive pacing therapy intervention consisting of occupational therapists helping patients to plan and pace activities utilising activity diaries to identify activities associated with fatigue and staying within their energy envelope [32]. This study incorporated standard medical care, cognitive behavioural therapy (CBT) and graded exercise therapy (GET) as comparator groups [32]. It is worth noting that the pacing group and the CBT group were both ‘encouraged’ to increase physical activity levels as long as participants did not exceed their energy envelope. Although not all five intervention studies explicitly mentioned the “Energy Envelope Theory”, which dictates that people with ME/CFS should not necessarily increase or decrease their activity levels, but moderate activity and practice energy conservation [59], all intervention studies used language analogous to this theory, such as participants staying within limits, within capacity, or similar.

The interventions included in this review were of varying durations, from a single 30-minute education session [46], a 3-week (one session a week) educational programme [48], a 3-week (3 x 60-90 minute sessions/week) educational programme [45], a 6-week rehabilitation programme [47], to a 24-week programme [32]. Intervention follow-up durations also varied across studies from immediately after [46], 1-week [48], 3-weeks [45], 3-months [47], and 1-year post-intervention [32].

#### 3.3.2 Observational studies of pacing

Eight studies were observational and, therefore, included no intervention. Observational study sample sizes ranged from 16 in a cross-sectional interview study[22] to 1428 in a cross-sectional survey[49]. One study involved a retrospective analysis of participants’ own pacing strategies varying from self-guided pacing or pacing administered by a therapist compared with implementation of CBT and GET [49]. Five involved a cross-sectional analysis of participants own pacing strategies which varied from activity adjustment, planning and acceptance [47,52], and the Energy Envelope method [55,57]. Two studies were prospective observational studies investigating the Energy Envelope theory [50,51]. Four studies [53,54,56,58] included in this review involved sub-analysis of results of the PACE trial [32].

### 3.4 Outcome measures

#### 3.4.1 Quantitative health outcomes

ME/CFS severity and general health status were the most common outcome measures across studies (16/17)[32,45–58,60]. Studies utilised different instruments, including the Short-Form 36 (SF-36; 8/16)[32,48,50,51,53–55,57], SF-12 (2/16)[47,60], ME symptom and illness severity (2/16)[49,52], Patient health (PHQ-15; 1/16)[56], DePaul symptom questionnaire (DSQ; 1/16)[55], and the Patient health questionnaire-9 (1/16)[47]. Additionally, some studies used diagnostic criteria for ME/CFS as an outcome measure to determine recovery [54,56,58].

Pain was assessed by most included studies (11/17) [32,46–48,50–52,54,56–58,60]. Two studies [56,58] included the international CDC criteria for CFS which contain five painful symptoms central to a diagnosis of CFS: muscle pain and joint pain. Other methods of assessment included Brief Pain Inventory (1/11)[50], Chronic Pain Coping Inventory (CPCI; 1/11)[46], Pain Self Efficacy Questionnaire (PSEQ; 1/11)[47], Tampa Scale for Kinesiophobia–version CFS (1/11)[46], algometry (1/11)[46], Knowledge of Neurophysiology of Pain Test (1/12)[46], Pain Catastrophizing Scale (1/11)[46], Pain Anxiety Symptoms Scale short version (PASS-20; 1/11)[47], Pain Numerical Rating Scale (NRS; 1/11)[60].

Fatigue or post-exertional malaise was assessed by 11 of the 17 studies [32,45,47,48,50,51,53,54,57,58,60]. Again, measurement instruments were divergent between studies and included the Chalder Fatigue Questionnaire (CFQ; 4/11) [32,47,54,60], Fatigue Severity Scale (2/11)[50,57], the Chronic Fatigue Syndrome Medical Questionnaire (1/11)[57], and Checklist Individual Strength (CIS; 2/11)[45,48].

Anxiety and depression were also common outcome measures, utilised by four studies (4/17)[47,50,56,60]. These were also assessed using different instruments including Hospital Anxiety and Depression Scale (HADS; 2/4)[56,60], Generalised Anxiety Disorder Assessment (1/4[47]), Beck Depression Inventory (BDI-II; 1/4)[50], Beck Anxiety Inventory (BAI; 1/4)[50], and Perceived Stress Scale (PSS; 1/4)[50].

Outcome measures also included sleep (2/17)[50,56], assessed by The Pittsburgh Sleep Quality Index (1/2)[50] and Jenkins sleep scale (1/2)[56]; and quality of life (2/17)[47,50] as assessed by the EuroQol five-dimensions, five-levels (EQ-5D-5L; 1/2)[47] and The Quality-of-Life Scale (1/2)[50]. Self-Efficacy was measured in four studies[47,50,56,57], assessed by the Brief Coping Orientation to Problems Experienced Scale (bCOPE; 1/4)[57] and the Chronic Disease Self-Efficacy measure (3/4)[47,50,56].

#### 3.4.2 Quantitative evaluation of pacing

Some studies (4/17)[22,47,49,60] included assessments of the participants’ experiences of pacing, using the Activity Pacing Questionnaire (APQ-28; 1/4[47], APQ-38 (2/4)[22,60]), a re-analysis of the 228 question survey regarding treatment (1/4) [49] originally produced by the ME Association [52], and qualitative semi-structured telephone interviews regarding appropriateness of courses in relation to individual patient needs (1/4)[22]. The APQ-28 and - 38 have been previously validated, but the 228 question survey has not. When outcome measures included physical activity levels (4/17), the Canadian Occupational Performance Measure (COPM) was used in two studies[45,48] and two studies used accelerometers to record physical activity [48,51]. Of these two studies, Nijs [48] examined accelerometery after a 3 week intervention based on the Energy Envelope Theory and Brown et al. [51] evaluated the Energy Envelope Theory of pacing over 12 months.

#### 3.4.3 Other outcomes

Two[50,56] of the 17 studies included structured clinical interviews for the Diagnostic and Statistical Manual of Mental Disorders, 4^th^ edition (DSM-IV) to assess psychiatric comorbidity and psychiatric exclusions. One study included a disability benefits questionnaire[52], and one study included employment and education questionnaire[52]. Additionally, satisfaction of primary care was also used as an outcome measure (2/17)[22,52] assessed using the Chronic Pain Coping Inventory (CPCI).

### 3.5 Efficacy of pacing interventions

The majority of studies (12/17)[22,45,47–53,55,57,60] highlighted improvements in at least one outcome following pacing (figure 2). When the effect of pacing was assessed by ME symptomology and general health outcomes, studies reported pacing to be beneficial[22,47,48,50–53,55]. It is worth noting however that pacing reportedly worsened ME symptoms in 14% of survey respondents, whilst improving symptoms in 44% of respondents [49]. Most studies using fatigue as an outcome measure reported pacing to be efficacious (7/10)[47,48,50,51,53,57,60]. However, one study reported no change in fatigue with a pacing intervention (1/10)[32], and 2/10 studies[50,60] reported a worsening of fatigue with pacing. Physical function was used to determine the efficacy of pacing in 11 studies[32,45,47,48,50,51,53,55–57,60]. Of these, the majority found pacing improved physical functioning (8/10)[45,47,48,50,51,53,55,57], with 1/10 [32] studies reporting no change in physical functioning, and 1/10 [56] reporting a worsening of physical functioning from pre- to post-pacing. Of the seven studies[32,46–48,50,51,57] which used pain to assess pacing efficacy, 4/7[47,48,50,57] reported improvements in pain and 3/7[32,48,50] reported no change in pain scores with pacing. All studies reporting quality of life (1/1)[50], self-efficacy (3/3)[47,50,56], sleep (2/2)[50,56], and depression and anxiety (4/4)[47,50,56,60], found pacing to be efficacious for ME/CFS participants.

**Fig. 2.**
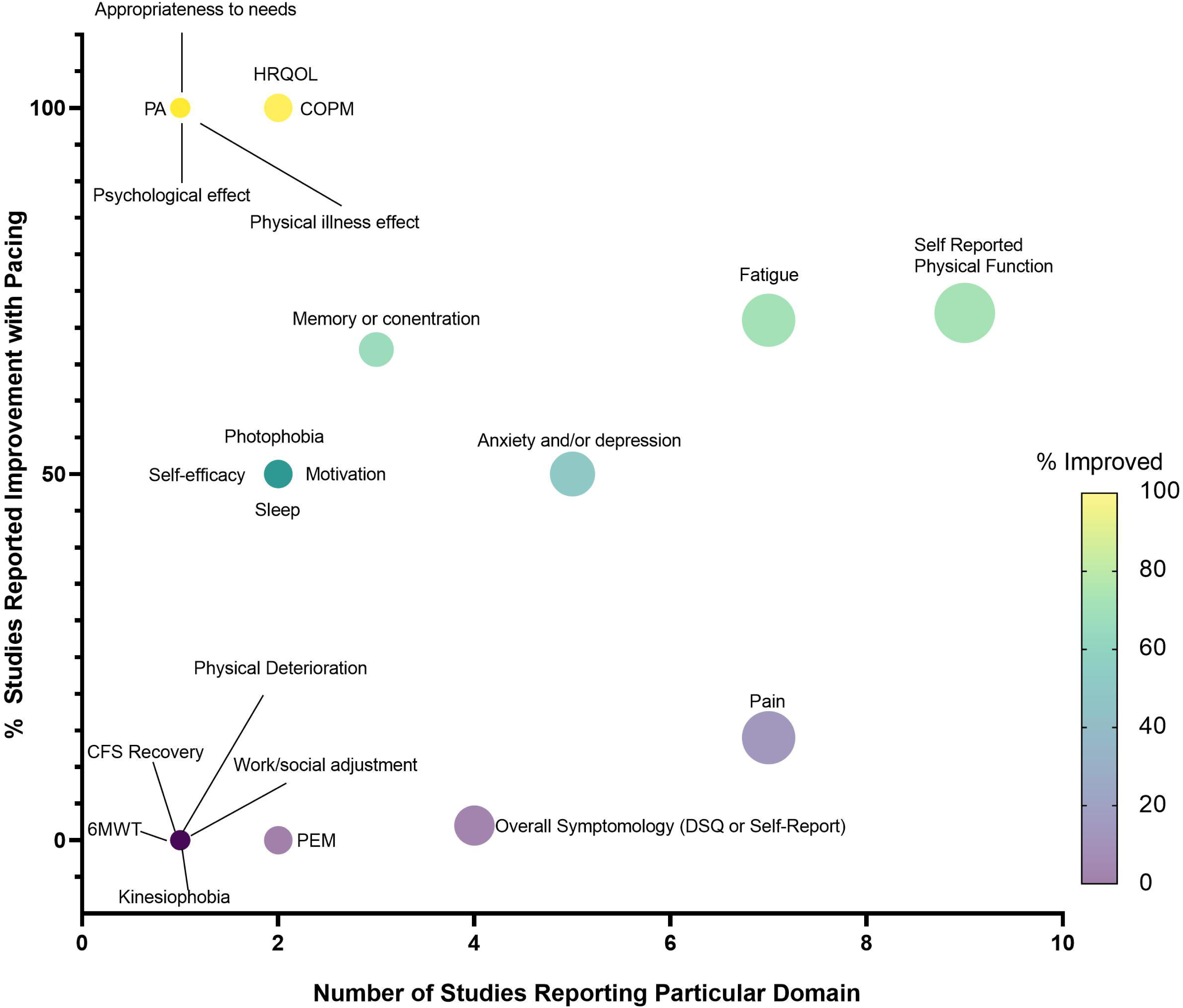
Bubble plot displaying number of studies reporting each domain (x-axis) and the percentage of studies reporting improvement with pacing (y-axis), including a coloured scale of improvement from 0-100%. PEM = post-exertional malaise, 6MWT = 6-minute walk time, CFS = chronic fatigue syndrome, DSQ = DePaul Symptom Questionnaire, PA = Physical Activity, HRQOL = Health-related quality of life, COPM = The Canadian Occupational Performance Measure.

### 3.6 Participant characteristics

The majority of studies (10/17)[22,47,49–51,55–58,60] did not report age of the participants. For those which did report age, this ranged from 32 ± 14 to 43 ± 13 years. Where studies reported sex (11/17)[32,45–48,51–55,57], this was predominantly female, ranging from 75% to 100% female. Only six studies[32,51,53–55,57] reported ethnicity, with cohorts predominantly Caucasian (94-98%). Time since diagnosis was mostly unreported (12/17)[22,45–47,49–51,55–58,60] but ranged from 32 to 96 months, with a cross-sectional survey reporting 2% of the participants were diagnosed 1-2 years previously; 6% 3-4 years since diagnosis; 13% 3-4 years since diagnosis; 12% 5-6 years since diagnosis; 20% 7-10 years since diagnosis; 29% 11-21 years since diagnosis; 13% 21-30 years since diagnosis; and 5% >30 years since diagnosis. Of the studies which reported comorbidities of the participants (6/17) [22,32,47,53,54,60], the comorbidities were chronic pain, depressive disorder, psychiatric disorder.

### 3.7 Study location

Of the 17 studies, 14 were from Europe[22,32,45–49,52–56,58,60], and three from North America[50,51,57]. Of the 14 studies[22,32,45–49,52–56,58,60] from Europe, ten[22,32,47,49,52–56,58,60] were conducted in the United Kingdom, three in Belgium[45,46,48], and one was a multicentred study between the United Kingdom and Norway[55].

### 3.8 Recruitment strategy

Of the 17 studies, three[50,51,57] used announcements in a newspaper and physician referrals to recruit participants, two[47,60] recruited patients referred by a consultant from a National Health Service (NHS) Trust following a pain diagnosis, two[49,52] concerned online platforms on the web, two[56,58] recruited from secondary care clinics, and two used the PACE trial databases[53,54]. Moreover, one study recruited from the hospital [55], one from physiotherapist referrals [22], two from specialist clinic centres [32,61], one from waiting list of rehabilitation centre [45], and one from medical files [46].

### 3.9 Study settings

Ten studies were carried out in hospital and clinic setting[22,32,45–48,55,56,58,60]. Two studies were performed on online platforms [49,52]. Three studies did not report study setting[50,51,57]. Two studies generated output from PACE trial databases [53,54]

### 3.10 Adherence and feasibility

All five intervention studies reported adherence rates (which they defined as number of sessions attended), which ranged from 4-44% (4% [46], 8% [32], 25% [45], 29% [48], and 44% [47]). One study reported the median number of rehabilitation programme sessions attended was five out of six possible sessions, with 58.9% [47] participants attending ≥5 sessions; 83.2% participants attending at least one educational session on activity pacing and 56.1% attending both activity pacing sessions.

## 4 Discussion

This scoping review summarises the existing literature, with a view to aid physicians and healthcare practitioners better summarise evidence for pacing in ME/CFS and use this knowledge for other post-viral fatiguing conditions. Overall, studies generally reported pacing to be beneficial for people with ME/CFS. The exception to this trend is the controversial PACE trial [33–36], which we will expand on in subsequent sections. We believe information generated within this review can facilitate discussion of research opportunities and issues that need to be addressed in future studies concerning pacing, particularly given the immediate public health issue of the long COVID pandemic. As mentioned, we found some preliminary evidence for improved symptoms following pacing interventions or strategies. However, we wish to caution the reader that the current evidence base is extremely limited and hampered by several limitations which preclude clear conclusions on the efficacy of pacing. Firstly, studies were of poor to fair methodological quality (indicated by the PEDro scores), often with small sample sizes, and therefore unknown power to detect change. Moreover, very few studies *implemented* pacing, with most studies merely consulting on people’s views on pacing. This may of course lead to multiple biases such as reporting, recruitment, survivorship, confirmation, availability heuristic, to name but a few. Thus, there is a pressing need for more high-quality interventions studies. Secondly, the reporting of pacing strategies used was inconsistent and lacked detail, making it difficult to describe current approaches, or implement them in future research or symptom management strategies. Furthermore, outcome evaluations varied greatly between studies. This prevents any appropriate synthesis of research findings.

The lack of evidence concerning pacing is concerning given pacing is the only NICE recommended management strategy for ME/CFS following the 2021 update [31]. Given the analogous nature of long COVID with ME/CFS, patients and practitioners will be looking to the ME/CFS literature for guidance for symptom management. There is an urgent need for high quality studies (such as RCTs) investigating the effectiveness of pacing and better reporting of pacing intervention strategies so that clear recommendations can be made to patients. If this does not happen soon, there will be serious healthcare and economic implications for years to come [62,63].

### 4.1 Efficacy of pacing

Most studies (12/17) highlighted improvements in at least one outcome measure following pacing. Pacing was self-reported to be the most efficacious, safe, acceptable, and preferred form of activity management for people with ME/CFS [52]. Pacing was reported to improve symptoms and improve general health outcomes [22,47,49,55,60], fatigue and PEM [45,47,48,50–53,57,60], physical functioning [45,47,48,50,53,55,57,60], pain [22,47,52,60], quality of life [47], self-efficacy [47,50], sleep [50,52], and depression and anxiety [47,50,60]. These positive findings provide hope for those with ME/CFS, and other chronic fatiguing conditions such as long COVID, to improve quality of life through symptom management.

Conversely, some studies reported no effects of pacing on ME/CFS symptoms [49], fatigue, physical functioning [32], or pain scores [46,58]. Some studies even found pacing to have detrimental effects in those with ME/CFS, including a worsening of symptoms in 14% of survey participants recalling previous pacing experiences [49]. Furthermore, a worsening of fatigue [32,56], and physical functioning from pre- to post-pacing [32,54,56,58] was reported by the PACE trial and sub-analysis of the PACE trial [53,54,58]. The PACE trial [32], a large RCT (n=639) comparing pacing with CBT and GET, reported GET and CBT were more effective for reducing ME/CFS-related fatigue and improving physical functioning than pacing. However, the methodology and conclusions from the PACE trial have been heavily criticised, mainly due to the authors lowering the thresholds they used to determine improvement [33–35,64]. With this in mind, Sharpe et al. [53] surveyed 75% of the participants from the PACE trial 1-year post-intervention and reported pacing improved fatigue and physical functioning, with effects similar to CBT and GET.

### 4.2 Lessons for pacing implementation

All pacing intervention studies (5/5) implemented educational or coaching sessions. These educational components were generally poorly reported in terms of the specific content and how and where they had been developed, with unclear pedagogical approaches. Consequently, even where interventions reported reduction in PEM or improved symptoms, it would be impossible to transfer that research into practice, future studies, or clinical guidance, given the ambiguity of reporting. Sessions typically contained themes of pacing such as activity adjustment (decrease, break-up, and reschedule activities based on energy levels), activity consistency (maintaining a consistently low level of activity to prevent PEM), activity planning (planning activities and rest around available energy levels), and activity progression (slowly progressing activity once maintaining a steady baseline) [32,45–48]. We feel it is pertinent to note here that although activity progression has been incorporated as a pacing strategy in these included studies, some view activity progression as a form of GET. The NICE definition of GET is “first establishing an individual’s baseline of achievable exercise or physical activity, then making fixed incremental increases in the time spent being physically active” [31]. Thus, this form of pacing can also be considered a type of ‘long-term GET’ in which physical activity progression is performed over weeks or months with fixed incremental increases in time spent being physically.

Intervention studies attempted to create behaviour change, through educational programmes to modify physical activity, and plan behaviours. However, none of these studies detailed integrating any evidence-based theories of behaviour change [65] or reported using any frameworks to support behaviour change objectives. This is unfortunate since there is good evidence that theory-driven behaviour change interventions result in greater intervention effects [66]. Indeed, there is a large body of work regarding methods of behaviour change covering public health messaging, education, and intervention design, which has largely been ignored by the pacing literature. Interventions relied on subjective pacing (5/5 studies), with strategies including keeping an activity diary (3/5 studies) to identify links between activity and fatigue [32,45,47]. Given the high prevalence of ‘brain fog’ within ME/CFS [67–70], recall may be extremely difficult and there is significant potential for under-reporting. Other strategies included simply asking participants to estimate energy levels available for daily activities (2/5 studies [45,48]). Again, this is subjective and relies on participants’ ability to recall previous consequences of the activity. Other methods of activity tracking and measuring energy availability, such as wearable technology [71–75] could provide a more objective measure of adherence and pacing strategy fidelity in future studies. Despite technology such as accelerometers being widely accessible since well-before the earliest interventional study included in this review (which was published in 2009 ^2^

), none of the interventional studies utilised objective activity tracking to track pacing and provide feedback to participants. One study considered accelerometery alongside an activity diary [48]. However, accelerometery was considered the outcome variable, to assess change in activity levels from pre- to post-intervention and was not part of the intervention itself (which was one pacing coaching sessions per week for 3 weeks). Moreover, most research-grade accelerometers cannot be used as part of the intervention since they have no ability to provide continuous feedback and must be retrieved by the research team in order to access any data. Consequently, their use is mostly limited to outcome assessments only. As pacing comprises a limit to physical activity to prevent push-crash cycles, it is an astonishing observation from this scoping review that only two studies objectively measured physical activity to quantify changes to activity as a result of pacing [48,51]. If the aim of pacing is to reduce physical activity, or reduce variations in physical activity (i.e., push-crash cycles), only two studies have objectively quantified the effect pacing had on physical activity, so it is unclear whether pacing was successfully implemented in any of the other studies.

By exploring the pacing strategies previously used, in both intervention studies and more exploratory studies, we can identify and recommend approaches to improve symptoms of ME/CFS. These approaches can be categorised as follows: activity planning, activity consistency, activity progression, activity adjustment and staying within the Energy Envelope [47,50,57,60]. Activity planning was identified as a particularly effective therapeutic strategy, resulting in improvement of mean scores of all symptoms included in the APQ-28, reducing current pain, improvement of physical fatigue, mental fatigue, self-efficacy, quality of life, and mental and physical functioning [47]. Activity planning aligns with the self-regulatory behaviour change technique ‘Action Planning’ [76] which is commonly used to increase physical activity behaviour. In the case of ME/CFS, activity planning is successfully used to minimise rather than increase physical activity bouts to prevent expending too much energy and avoid PEM. Activity consistency, meaning undertaking similar amounts of activity each day, was also associated with reduced levels of depression, exercise avoidance, and higher levels of physical function [60]. Activity progression was associated with higher levels of current pain. Activity adjustment associated with depression and avoidance, and lower levels of physical function [60]. Staying within the Energy Envelope was reported to reduce PEM severity [50,57], improve physical functioning [50,57] and ME/CFS symptom scores [50], and more hours engaged in activity than individuals with lower available energy [50]. These results suggest that effective pacing strategies would include activity planning, consistency, and energy management techniques while avoiding progression. This data is, of course, limited by the small number of mostly low-quality studies and should be interpreted with some caution. Nevertheless, these are considerations that repeatedly appear in the literature and, as such, warrant deeper investigation. In addition, and as outlined earlier, most studies are relatively old, and we urgently need better insight into how modern technologies, particularly longitudinal activity tracking and contemporaneous heart-rate feedback, might improve (or otherwise) adaptive pacing. Such longitudinal tracking would also enable activities and other behaviours (sleep, diet, stress) to be linked to bouts of PEM. Linking would enable a deeper insight into potential PEM triggers and mitigations that might be possible.

### 4.3 The PACE Trial

We feel it would be remiss of us to not specifically address the PACE trial within this manuscript, as five of the 17 included studies resulted from the PACE trial [32,53,54,56,58]. There has been considerable discussion around the PACE trial, which has been particularly divisive and controversial [34–36,56,64,77,78]. In the PACE trial, GET and CBT were deemed superior to pacing by the authors. Despite its size and funding, the PACE trial has received several published criticisms and rebuttals. Notably, NICE’s most recent ME/CFS guideline update removed GET and CBT as suggested treatment options, which hitherto had been underpinned by the PACE findings. While we will not restate the criticisms and rebuttals here, what is not in doubt, is that the PACE trial has dominated discussions of pacing, representing almost a third of all the studies in this review. However, the trial results were published over a decade ago, with the study protocol devised almost two decades ago [79]. The intervening time has seen a revolution in the development of mobile and wearable technology and an ability to remotely track activity and provide real-time feedback in a way which was not available at that time. Furthermore, there has been no substantive research since the PACE trial that has attempted such work. Indeed, possibly driven by the reported lack of effect of pacing in the PACE trial, this review has demonstrated the dearth of progress and innovation in pacing research since its publication. Therefore, regardless of its findings or criticisms, the pacing implementation in the PACE trial is dated, and there is an urgent need for more technologically informed approaches to pacing research.

### 4.4 Limitations of the current evidence

The first limitation to the literature included in this scoping review is that not all studies followed the minimum data set (MDS) of patient-reported outcome measures (PROMs) agreed upon by the British Association of CFS/ME Professionals (BACME) (fatigue, sleep quality, self-efficacy, pain/discomfort, anxiety/depression, mobility, activities of daily living, self-care, and illness severity) [80,81]. All but one study included in this review measured illness severity, most studies included fatigue and pain/discomfort, and some studies included assessments of anxiety/depression. There was a lack of quantitative assessment of sleep quality, self-efficacy, mobility, activities of daily living, and self-care. Therefore, studies did not consistently capture the diverse nature of the symptoms experienced, with crucial domains missing from the analyses. The MDS of PROMs were established in 2012 [80,81] and therefore, for studies published out prior to 2012, these are not applicable [32,46,48,50,51]. However, for the 12 studies carried out after this time, the MDS should have been considered elucidate the effects of pacing on ME/CFS. Importantly, despite PEM being a central characteristic of ME/CFS, only two studies included PEM as an outcome measure [52,57]. This may be because of the difficulty of accurately measuring fluctuating symptoms, as PEM occurs multiple times over a period of months, and therefore pre- to post- studies and cross-sectional designs cannot adequately capture PEM incidence. Therefore, it is likely studies opted for measuring general fatigue instead. More appropriate longitudinal study designs are required to track PEM over time to capture a more representative picture of PEM patterns. Secondly, reporting of participant characteristics was inadequate, but in the studies that did describe participants, characteristics were congruent with the epidemiological literature and reporting of ME/CFS populations (i.e., 60-65% female) [82]. Therefore, in this respect, studies included herein were representative samples. However, the lack of reporting of participant characteristics limits inferences we can draw concerning any population-related effects (i.e. whether older, or male, or European, or people referred by a national health service would be more or less likely to respond positively to pacing). Thirdly, comparison groups (where included) were not ideal, with CBT or GET sometimes used as comparators to pacing [32], and often no true control group included. Penultimately, there is a distinct lack of high-quality RCTs (as mentioned throughout this manuscript). Finally, in reference to the previous section, inferences from the literature are dated and do not reflect the technological capabilities of 2023.

### 4.5 Recommendations for advancement of the investigative area

It is clear from the studies included in this scoping review for the last decade or more, progress and innovation in pacing research have been limited. This is unfortunate for several reasons. People with ME/CFS or long COVID are, of course, invested in their recovery. From our patient and public involvement (PPI) group engagement, it is clear many are ahead of the research and are using wearable technology to track steps, heart rate, and, in some cases, heart rate variability to improve their own pacing practice. While the lack of progress in the research means this is an understandable response by patients, it is also problematic. Without underpinning research, patients may make decisions based on an individual report of trial-and-error approaches given the lack of evidence-based guidance.

A more technologically-informed pacing approach could be implemented by integrating wearable trackers [74,75,86,87] to provide participants with live updates on their activity and could be integrated with research-informed messaging aimed at supporting behaviour change, as has been trialled in other research areas [88–91]. However, more work is needed to evaluate how to incorporate wearable activity trackers and which metrics are most helpful.

A more technologically-informed approach could also be beneficial for longitudinal symptom tracking, particularly useful given the highly variable symptom loads of ME/CFS and episodic nature of PEM. This would overcome reliance on assessments at a single point in time (as the studies within this review conducted). Similarly, mobile health (mHealth) approaches also allow questionnaires to be digitised to make it easier for participants to complete if they find holding a pen or reading small font problematic [92]. Reminders and notifications can also be helpful for patients completing tasks [74,93–95]. This approach has the added advantage of allowing contemporaneous data collection rather than relying on pre- to post-intervention designs limited by recall bias. Future work must try to leverage these approaches, as unless we collect large data sets on symptoms and behaviours (i.e. activity, diet, sleep, and pharmacology) in people with conditions like ME/CFS we will not be able to leverage emerging technologies such as AI and machine learning to improve the support and care for people with these debilitating conditions. The key areas for research outline in the NICE guidelines (2021 update) speaks to this, with specific mention of improved self-monitoring strategies, sleep strategies, and dietary strategies, all of which can be measured using mHealth approaches, in a scalable and labour-inexpensive way.

### 4.6 The potential for existing pacing research to address the long COVID pandemic

There is now an urgent public health need to address long COVID, with over 200 million sufferers worldwide [27]. Given the analogous symptomology between ME/CFS and long COVID, and the lack of promising treatment and management strategies in ME/CFS, pacing remains the only strategy for managing long COVID symptoms. This is concerning as the quality of evidence to support pacing is lacking. Given long COVID has reached pandemic proportions, scalable solutions will be required. In this context, we propose that technology should be harnessed to a) deliver, but also b) evaluate, pacing. We recently reported on a just- in-time adaptive intervention to *increase* physical activity during the pandemic [75]. However, this method could be adapted to *decrease* or maintain physical activity levels (i.e., pacing) in long COVID. This method has the advantage of scalability and remote data collection, reducing resource commitments and participant burden, essential for addressing a condition with so many sufferers.

## 5 Conclusion

This review highlights the need for more studies concerning pacing in chronic fatiguing conditions. Future studies would benefit from examining pacing’s effect on symptomology and PEM with objectively quantified pacing, over a longer duration of examination, using the MDS. It is essential this is conducted as an RCT, given that in the case of long COVID, participants may improve their health over time, and it is necessary to determine whether pacing exerts an additional effect over time elapsing. Future studies would benefit from digitising pacing to support individuals with varying symptom severity and personalise support. This would improve accessibility and reduce selection bias, in addition to improving scalability of interventions. Finally, clinicians and practitioners should be cognisant of the strength of evidence reported in this review and should exert caution when promoting pacing in their patients, given the varying methods utilised herein.

## Data Availability

All data produced in the present work are contained in the manuscript

## List of Abbreviations

APQ: Activity Pacing Questionnaire
BAI: Beck Anxiety Inventory
BDI-II: Beck Depression Inventory
bCOPE: Brief Coping Orientation to Problems Experienced Scale
COPM: Canadian Occupational Performance Measure
CDC: Centers for disease control and prevention
CFQ: Chalder Fatigue Questionnaire
CIS: Checklist Individual Strength
CPCI: Chronic Pain Coping Inventory
CBT: Cognitive behavioural therapy
CENTRAL: Cochrane Central Register of Controlled Trials
DSQ: DePaul symptom questionnaire
EQ-5D-5L: EuroQol five-dimensions, five-levels questionnaire
GET: Graded exercise therapy
HADS: Hospital Anxiety and Depression Scale
ME/CFS: Myalgic encephalomyelitis/chronic fatigue syndrome
PSEQ: Pain Self Efficacy Questionnaire
PASS-20: Pain Anxiety Symptoms Scale short version
NRS: Pain Numerical Rating Scale
PHQ: Patient health questionnaire
PROMs: Ppatient reported outcome measures
PEDro: Physiotherapy Evidence Database
PSS: Perceived Stress Scale
PEM: Post exertional malaise
PRISMA-ScR: Preferred Reporting Items for Systematic Reviews and Meta-Analyses extension for scoping reviews
RCT: Randomised control trial
SF: Short-Form

## Declarations

### Ethical approval and content to participate

This manuscript did not involve human participants, data, or tissues, so did not require ethical approval.

### Consent for publication

This paper does not contain any individual person’s data in any form.

### Availability of data and materials

The datasets used and/or analysed during the current study are available from the corresponding author on reasonable request.

### Competing interests

We report no financial and non-financial competing interests.

### Funding

This work was supported by grants from the National Institute for Health and Care Research (COV-LT2-0010) and the funder had no role in the conceptualisation, design, data collection, analysis, decision to publish, or preparation of the manuscript.

### Authors’ contributions

Authors’ contributions are given according to the CRediT taxonomy as follows: Conceptualization, N.E.M.S-H., M.M., L.D.H, and N.F.S.; methodology, N.E.M.S-H., M.M., L.D.H., and N.F.S.; software, N.E.M.S-H., M.M., L.D.H., and N.F.S.B.; validation, N.E.M.S-H., M.M., L.D.H, and N.F.S.; formal analysis, N.E.M.S-H., M.M., L.D.H., and N.F.S.; investigation, N.E.M.S-H., M.M., L.D.H., and N.F.S.; resources, L.D.H., J.O., D.C., N.H., J.L.M., and N.F.S.; data curation, N.E.M.S.-H., M.M., L.D.H., and N.F.S.; writing—original draft preparation, N.E.M.S.-H., M.M., L.D.H., and N.F.S.; writing—review and editing, N.E.M.S-H., M.M., L.D.H., J.O., D.C., N.H., R.M., J.L.M., J.I., and N.F.S.; visualisation, N.E.M.S-H. and M.M., supervision, N.F.S; project administration, N.E.M.S-H., M.M., L.D.H., and N.F.S.; funding acquisition, L.D.H., J.O., D.C., N.H., J.L.M., J.I., and N.F.S. All authors have read and agreed to the published version of the manuscript.

## Acknowledgements

We have no acknowledgements to make.

## Notes

### Competing Interest Statement

The authors have declared no competing interest.

