## appendix 1 for "A Scoping Review of ‘Pacing’ for Management of Myalgic Encephalomyelitis/Chronic Fatigue Syndrome (ME/CFS): Lessons Learned for the Long COVID Pandemic"

**Appendix 1. Full Search Strings**

‘ME/CFS’ OR ‘ME’ OR ‘CFS’ OR ‘chronic fatigue syndrome’ OR ‘PEM’ OR ‘post exertional malaise’ OR ‘pene’ OR ‘post-exertion neurogenic exhaust’ AND ‘pacing’

‘ME/CFS’ OR ‘ME’ OR ‘CFS’ OR ‘chronic fatigue syndrome’ OR ‘PEM’ OR ‘post exertional malaise’ OR ‘pene’ OR ‘post-exertion neurogenic exhaust’ AND ‘adaptive pacing’.

‘Myalgic encephalomyelitis/chronic fatigue syndrome’ OR ‘Myalgic encephalomyelitis’ OR ‘chronic fatigue syndrome’ OR ‘PEM’ OR ‘post exertional malaise’ OR ‘pene’ OR ‘post-exertion neurogenic exhaust’ AND ‘pacing’

‘Myalgic encephalomyelitis/chronic fatigue syndrome’ OR ‘Myalgic encephalomyelitis’ OR ‘CFS’ OR ‘chronic fatigue syndrome’ OR ‘PEM’ OR ‘post exertional malaise’ OR ‘pene’ OR ‘post-exertion neurogenic exhaust’ AND ‘adaptive pacing’.
